# Predicting Continuous Cognitive Decline: The Generalizability of a Multimodal Machine Learning Approach Including Structural MRI and Non-Brain Data

**DOI:** 10.64898/2026.01.13.26343927

**Authors:** Roya Melanie Hüppi, Nicolas Langer, Bruno Hebling Vieira, the Alzheimer’s Disease Neuroimaging Initiative

**Author notes:** Corresponding Author: Bruno Hebling Vieira. Data used in preparation of this article were obtained from the Alzheimer’s Disease Neuroimaging Initiative (ADNI) database (adni.loni.usc.edu). As such, the investigators within the ADNI contributed to the design and implementation of ADNI and/or provided data but did not participate in analysis or writing of this report. A complete listing of ADNI investigators can be found at: http://adni.loni.usc.edu/wp-content/uploads/how_to_apply/ADNI_Acknowledgement_List.pdf.

## Abstract

Aging is often accompanied by cognitive decline, but the extent, timing, and severity of this process is subject to large inter-individual variability. Predicting cognitive decline along a continuum, encompassing healthy age-related decline, mild cognitive impairment, and dementia, allows for more precise individual-level predictions. Previous work has demonstrated that machine learning (ML) models combining risk factors, clinical, neuropsychological, and structural magnetic resonance imaging (MRI) data can predict continuous cognitive decline. However, generalizability across independent datasets has rarely been evaluated. This study aimed to replicate previous findings using an independent dataset and to assess the models’ generalizability across acquisition sites. Multi-target random forest regression models were employed to predict annual rates of decline of the Clinical Dementia Rating Scale Sum of Boxes (CDR-SOB) and the Mini-Mental State Examination (MMSE). Results showed that adding structural MRI data to non-brain data enhanced model performance in the large-scale novel ADNI (*N* = 1237) cohort, as previously attested for OASIS-3 (*N* = 662). Further, detectable reductions in performance were measured when models were tested across datasets compared to within datasets. This performance degradation was not explained by distributional shifts of the target variables. Additionally, models trained on the most important features of the respective training set achieved nearly identical performance as those trained on all features when tested externally, suggesting high redundancy among predictors. In summary, multimodal ML models predicting continuous cognitive decline generalize partially to unseen cohorts, with statistically significant performance degradation, whereas unimodal models trained on structural MRI features do not.

## 1 Introduction

Substantial healthcare, economic, and social costs are caused by pathological age-related cognitive decline (Scheltens et al., 2016; Scheltens et al., 2021; Zvěřová, 2019). Cognitive decline manifests, for instance, as deterioration of executive functioning, memory, or language (Plassman et al., 2010). It represents one of the criteria for dementia (McKhann et al., 2011), the worldwide prevalence of which was predicted to be about 50 million individuals in 2018 and is expected to more than triple to 152 million by the year 2050 (Alzheimer’s Disease International, 2018; Scheltens et al., 2021). The most common cause of dementia is Alzheimer’s disease (AD), one of the biggest healthcare challenges of this century (Alzheimer’s Disease International, 2018; Scheltens et al., 2016; Scheltens et al., 2021).

Biomarkers of cognitive impairment include indicators of brain atrophy from structural magnetic resonance imaging (MRI) (Jack et al., 2010). However, translating a potentially vast number of biomarkers into actionable insights is a complex and demanding task. Machine learning (ML) has been proposed as a technique to objectively translate the markers of brain atrophy and risk factors into diagnostic and prognostic predictions (Woo et al., 2017; Zhao et al., 2023). With ML, large datasets including imaging and genetic biomarkers as well as clinical, lifestyle and environmental data can be jointly analyzed (Fabrizio et al., 2021; Korolev et al., 2016).

ML approaches have provided promising results in predicting individuals’ progression from mild cognitive impairment (MCI) to AD (Cui et al., 2011; Davatzikos et al., 2011; Korolev et al., 2016; Moradi et al., 2015; Young et al., 2013) or predicting assignment to data-driven trajectory clusters of future cognitive decline (Bhagwat et al., 2018; Tam et al., 2022). However, the division into normal aging, MCI, and dementia and the diagnostic threshold between normal and pathological cognitive changes may reflect somewhat artificial boundaries (Dubois et al., 2021; Jack et al., 2000; Plassman et al., 2010). An alternative view suggests studying cognitive decline as a continuum, rather than adhering to discrete diagnostic categories, enabling a more accurate depiction of the underlying individual change in abilities and to facilitate a more detailed and robust prediction (Vieira et al., 2022).

Using data from the Open Access Series of Imaging Studies (OASIS-3), a large open-access data collection, Vieira et al. (2022) showed that combining structural MRI data and non-brain data, such as risk factors and neuropsychological test variables, improved performance of the prediction of yearly rates of future score change of the Clinical Dementia Rating Scale Sum of Boxes (CDR-SOB) and the Mini-Mental State Examination (MMSE). This result was in line with previous findings revealing that combining non-brain data with structural MRI data improved the prediction of dementia progression (Bhagwat et al., 2018; Korolev et al., 2016). However, the practical utility of such ML models depends on their generalizability to unseen, independently acquired data (Myszczynska et al., 2020; Woo et al., 2017). This generalizability can be hampered by heterogeneity arising from systematic differences between data acquisition sites, commonly known as site effects, e.g., different MRI acquisition parameters or study populations, which introduce bias (Chen et al., 2022). Yet, ML models’ generalizability is often not assessed.

Therefore, the present study used data from the large, publicly available Alzheimer’s Disease Neuroimaging Initiative (ADNI) dataset to investigate whether the inclusion of structural MRI data in the model significantly improves the prediction of continuous cognitive decline compared to models using only non-brain data. It further aimed to evaluate the generalizability of an ML model including non-brain data and structural MRI data for predicting future continuous cognitive decline across datasets. OASIS-3 data and ADNI data were used for this purpose. We tested whether the internal performance of a model predicting continuous cognitive decline within the ADNI dataset differed significantly from that of a model trained and evaluated within the OASIS-3 dataset. In addition, we assessed the difference in performance when predicting continuous cognitive decline within each dataset versus across the two independent datasets. We then evaluated how the models predicting across datasets would perform across different subgroups. In a post hoc analysis, we explored the potential of matching the distributions of the target variables of the test set to those of the respective training set to mitigate bias. Lastly, we compared the performance and generalizability of a model trained on the 15 most predictive features of the OASIS-3 dataset to the performance of a model trained on all non-brain features and structural MRI features of the OASIS-3 dataset.

## 2 Methods

### 2.1 Study Population and Session Selection

Data from two cohorts were included in this study. Relevant institutional ethics approvals were obtained in advance from the respective source institutions. The first dataset consisted of 662 participants from the OASIS-3 project^1^. OASIS-3 is a longitudinal, multimodal, open-access data collection stemming from various studies at the Washington University Knight Alzheimer Disease Research Center. Data from different types of sessions, ergo clinical (non-brain data describing personal characteristics, health, cognitive and everyday functioning) and neuropsychological sessions (non-brain data from neuropsychological tests) (see Non-Brain Data) as well as structural MRI sessions (see Structural MRI Data) were incorporated (LaMontagne et al., 2019; Vieira et al., 2022).

Baseline sessions (from clinical, neuropsychological, and MRI sessions) were used as input data, and follow-up clinical sessions, i.e., all clinical sessions after the baseline clinical sessions, were used as targets for the prediction of future continuous cognitive decline. To establish baseline data, MRI sessions were matched with their closest clinical session with a maximal absolute time difference of one year. Further, the closest neuropsychological session within one year of the MRI baseline session, when available, was selected as baseline data. However, baseline information from neuropsychological testing was regarded as optional and the lack thereof was not a criterion for exclusion (see Treatment of Missing Data for more information on how missing data was handled). No data prior to the selected baseline sessions were considered in the analysis. All baseline and follow-up clinical sessions were used to estimate the individual rate of the yearly cognitive decline. To ensure a reliable estimation, participants needed at least three clinical sessions, i.e., one baseline and 2 or more follow-up sessions where both MMSE and CDR-SOB were assessed. The session selection led to a sample of 662 participants from OASIS-3.

For the second dataset, data were obtained from the ADNI database (adni.loni.usc.edu). The ADNI is led by Principal Investigator Michael W. Weiner, MD, and was launched in 2003 as a public-private partnership. The primary objective of ADNI has been to test whether serial MRI, positron emission tomography (PET), other biological markers, and clinical and neuropsychological assessment could be used jointly to measure the progression of MCI and early AD. For up-to-date information, see www.adni-info.org. The procedure of the data selection was identical as described above for the OASIS-3 cohort. Consequently, 1237 ADNI participants that fulfilled inclusion criteria were included in this study (see Supplementary Material, Figure A). Sample characteristics of the OASIS-3 and ADNI datasets are shown in Table 1. Chi-squared tests were performed to test for differences between datasets concerning sex and diagnoses, and *t*-tests were calculated regarding education, age, and the slopes of CDR-SOB and MMSE (α = .05).

**Table 1.**
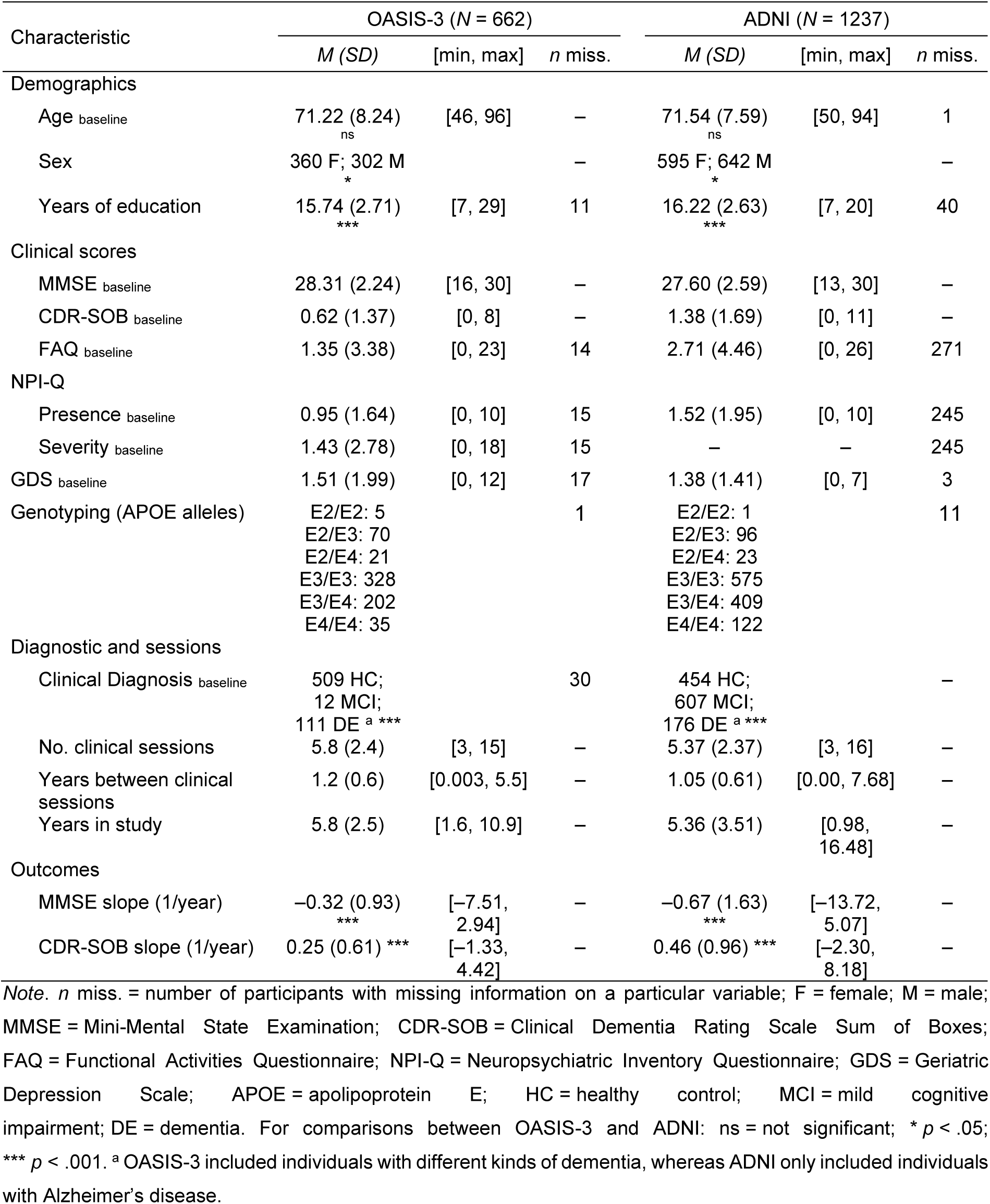
Sample Characteristics of the OASIS-3 and ADNI Datasets.

| Characteristic | OASIS-3 (N = 662) |  |  | ADNI (N = 1237) |  |  |
| --- | --- | --- | --- | --- | --- | --- |
|  | M (SD) | [min, max] | n miss. | M (SD) | [min, max] | n miss. |
| **Demographics** |  |  |  |  |  |  |
| Age baseline | 71.22 (8.24)<br>ns | [46, 96] | – | 71.54 (7.59)<br>ns | [50, 94] | 1 |
| Sex | 360 F; 302 M<br>* |  | – | 595 F; 642 M<br>* |  | – |
| Years of education | 15.74 (2.71)<br>*** | [7, 29] | 11 | 16.22 (2.63)<br>*** | [7, 20] | 40 |
| **Clinical scores** |  |  |  |  |  |  |
| MMSE baseline | 28.31 (2.24) | [16, 30] | – | 27.60 (2.59) | [13, 30] | – |
| CDR-SOB baseline | 0.62 (1.37) | [0, 8] | – | 1.38 (1.69) | [0, 11] | – |
| FAQ baseline | 1.35 (3.38) | [0, 23] | 14 | 2.71 (4.46) | [0, 26] | 271 |
| **NPI-Q** |  |  |  |  |  |  |
| Presence baseline | 0.95 (1.64) | [0, 10] | 15 | 1.52 (1.95) | [0, 10] | 245 |
| Severity baseline | 1.43 (2.78) | [0, 18] | 15 | – | – | 245 |
| GDS baseline | 1.51 (1.99) | [0, 12] | 17 | 1.38 (1.41) | [0, 7] | 3 |
| Genotyping (APOE alleles) | E2/E2: 5<br>E2/E3: 70<br>E2/E4: 21<br>E3/E3: 328<br>E3/E4: 202<br>E4/E4: 35 |  | 1 | E2/E2: 1<br>E2/E3: 96<br>E2/E4: 23<br>E3/E3: 575<br>E3/E4: 409<br>E4/E4: 122 |  | 11 |
| **Diagnostic and sessions** |  |  |  |  |  |  |
| Clinical Diagnosis baseline | 509 HC;<br>12 MCI;<br>111 DE<sup>a</sup> *** |  | 30 | 454 HC;<br>607 MCI;<br>176 DE<sup>a</sup> *** |  | – |
| No. clinical sessions | 5.8 (2.4) | [3, 15] | – | 5.37 (2.37) | [3, 16] | – |
| Years between clinical sessions | 1.2 (0.6) | [0.003, 5.5] | – | 1.05 (0.61) | [0.00, 7.68] | – |
| Years in study | 5.8 (2.5) | [1.6, 10.9] | – | 5.36 (3.51) | [0.98, 16.48] | – |
| **Outcomes** |  |  |  |  |  |  |
| MMSE slope (1/year) | –0.32 (0.93)<br>*** | [–7.51, 2.94] | – | –0.67 (1.63)<br>*** | [–13.72, 5.07] | – |
| CDR-SOB slope (1/year) | 0.25 (0.61) *** | [–1.33, 4.42] | – | 0.46 (0.96) *** | [–2.30, 8.18] | – |

Note. n miss. = number of participants with missing information on a particular variable; F = female; M = male; MMSE = Mini-Mental State Examination; CDR-SOB = Clinical Dementia Rating Scale Sum of Boxes; FAQ = Functional Activities Questionnaire; NPI-Q = Neuropsychiatric Inventory Questionnaire; GDS = Geriatric Depression Scale; APOE = apolipoprotein E; HC = healthy control; MCI = mild cognitive impairment; DE = dementia. For comparisons between OASIS-3 and ADNI: ns = not significant; \* $p < .05$ ; \*\*\* $p < .001$ . <sup>a</sup> OASIS-3 included individuals with different kinds of dementia, whereas ADNI only included individuals with Alzheimer's disease.

### 2.2 Modeling the Prediction of Continuous Cognitive Decline

For the prediction of the rate of future continuous cognitive decline, input data included non-brain data and structural MRI data. Non-brain data originally consisted of 66 features that entered the model as the *non-brain* modality (see Treatment of Missing Data). Specifically, three features with demographic information (i.e., age, sex, and education), 23 clinical assessment features, 18 neuropsychological features, three features concerning Apolipoprotein E (APOE) count, 17 features with health information, the cognitive diagnosis, and the number of clinical sessions preceding the baseline session were incorporated into the predictive model. For the *structural MRI* modality, structural MRI data contained 35 features that served as input data, consisting of 14 features regarding regional grey matter volume and 21 features concerning global or hemispheric measurements. Further details on the non-brain features and structural MRI features are provided in Non-Brain Data, Structural MRI Data, and the Supplementary Material, Table B.

Trajectories of two clinical assessments, CDR-SOB and MMSE, were estimated to quantify future continuous cognitive decline, following the approach of Vieira et al. (2022). For this, an ordinary least squares linear regression model was fitted for each participant and for each target variable separately:

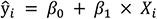

where ŷ_i_ is the estimated CDR-SOB or MMSE score of the follow-up clinical session, the intercept *β*_0_ = y_0_is the CDR-SOB or MMSE score at baseline and *X_i_* is the non-rounded number of years between the MRI session and the follow-up clinical session. *β*_1_is the resulting parameter which represents either the slope of CDR-SOB or MMSE change. Both the slopes of CDR-SOB and MMSE change were simultaneously the targets of the predictive analysis, as multi-output models can increase predictive accuracy when predicting across datasets, and therefore improve generalizability compared to single-output models (Rahim et al., 2017; Vieira et al., 2022).

### 2.3 Non-Brain Data

Non-brain data incorporated a broad spectrum of measures to describe personal characteristics at baseline. This includes demographics, such as sex, age, and education. Being a woman, advanced age, and having less education have been previously described as risk factors for AD (Altmann et al., 2014; Caamaño-Isorna et al., 2006; Chêne et al., 2015; Scheltens et al., 2021). Similar risk factors also exist for the less prevalent non-AD dementias (Rasmussen et al., 2018)

Furthermore, the non-brain data contained clinical scores, including the CDR-SOB (Morris, 1993), MMSE (Folstein et al., 1975), Functional Activities Questionnaire (FAQ; Jette et al., 1986), Geriatric Depression Scale (GDS; Yesavage et al., 1982) and Neuropsychiatric Inventory Questionnaire (NPI-Q; Kaufer et al., 2000). While the CDR-SOB and the MMSE are often used instruments for screening and to stage the severity of AD (Mitchell, 2013; O’Bryant et al., 2008), the FAQ is commonly used to assess instrumental activities of daily living, for example, paying bills or remembering appointments, and therefore to distinguish individuals with AD from individuals with MCI and healthy controls (HC) (Teng et al., 2010). In addition, FAQ scores were found to predict progression from MCI to AD in ADNI data (Berezuk et al., 2023). In line with research postulating neuropsychiatric symptoms, among them depressive symptoms, as risk factors for developing dementia (Acosta et al., 2018; Livingston et al., 2020), NPI-Q scores as well as GDS scores have shown predictive value for predicting conversion from MCI to AD (Dolcet-Negre et al., 2023; Mallo et al., 2020).

Moreover, neuropsychological scores resulting from the Boston Naming Test (BNT; Borod et al., 1980), Trail Making Test (TMT; Bucks, 2013), word fluency, and Wechsler Memory Scale-Revised (WMS-R; Elwood, 1991) were part of the non-brain data. In previous research, all neuropsychological tests included have been associated with cognitive decline. The BNT, a widely used confrontation naming test (Rabin et al., 2005; Williams et al., 1989), was demonstrated to be a predictor of conversion from MCI to AD (Eckerström et al., 2013). Likewise, the TMT, measuring processing speed, mental flexibility, sequencing, and visual-motor skills (Bowie & Harvey, 2006; Thompson et al., 1999), predicted conversion from MCI to AD (Eckerström et al., 2015; Ewers et al., 2012) and dementia progression (Edwin et al., 2021). Further, word fluency, a measurement involving executive functioning and language processing (Ghanavati et al., 2019; Whiteside et al., 2016), was used to distinguish individuals with MCI and AD (Fernaeus et al., 2008) as well as to predict the risk of incident MCI (Jacobs et al., 2021). Subtests of the WMS-R, providing an estimate of overall memory functioning (Elwood, 1991), were argued to be predictors for differentiating between individuals with dementia and healthy subjects (Derrer et al., 2001).

APOE ε2, ε3, and ε4 allele counts (being either 0, 1, or 2) were also incorporated in the non-brain data. Carrying at least one APOE ε4 allele is one of the strongest risk factors for AD, while APOE ε2 represents a potential protective genetic variant associated with a lower risk of AD compared with the neutral, and most prevalent, third variant, APOE ε3 (Scheltens et al., 2021; Singh et al., 2006; Wood et al., 2023). Evidence also implicates APOE in non-AD dementias, such as vascular cognitive impairment (Rasmussen et al., 2018) and dementia with Lewy’s bodies (Mirza et al., 2019).

Clinically conferred diagnostic classifications, either HC, MCI, or dementia, were included as part of the non-brain data features. Individuals with MCI have previously been found to demonstrate a higher rate of cognitive decline than HC, but a slower decline than individuals with mild AD in longitudinal data (R. C. Petersen et al., 1999). Accounting for the health information of the participants, measures regarding cardio-and cerebrovascular health, diabetes, and hypercholesterolemia were included as well, as they all constitute risk factors for AD (Livingston et al., 2020). Finally, to account for potential retest effects, the number of clinical sessions preceding the selected baseline session, i.e., sessions that could not be matched with an MRI session, was included as non-brain data.

### 2.4 Structural MRI Data Acquisition

Since the addition of structural MRI data to non-brain data has previously been shown to improve the performance of models predicting dementia progression (Bhagwat et al., 2018; Korolev et al., 2016) and continuous cognitive decline (Vieira et al., 2022), structural MRI data entered the model as input data. In the OASIS-3 study, MRI data were acquired on Siemens 3T scanners. The most predominantly used parameter combination was voxel size = 1 × 1 × 1 mm^3^, repetition time (TR) = 2400 ms, and echo time (TE) = 3 ms. In ADNI, 3T MRI data were acquired on scanners from three different vendors: Siemens, Philips, and GE HealthCare. The two most frequently used parameter combinations were voxel size = 1.2 × 1 × 1 mm^3^, TR = 2300 ms, and TE = 3 ms as well as voxel size = 1 × 1 × 1 mm^3^, TR = 2300 ms, and TE = 3 ms. 1.5T MRI scans from ADNI were not used. Further information about MRI data acquisition is displayed in the Supplementary Material, Table A.

### 2.5 MRI Preprocessing

Results reported in this study come from data preprocessed using *fMRIPrep* 1.4.1 in OASIS-3 and *sMRIPprep* 0.9.2 in ADNI (RRID: SCR_016216; Esteban et al., 2019; Esteban et al., 2023), which are based on *Nipype* 1.2.0 (OASIS-3) and *Nipype* 1.8.1 (ADNI) (RRID: SCR_002502; Esteban et al., 2022; Gorgolewski et al., 2011; Gorgolewski et al., 2019). Correction for intensity non-uniformity (INU) of T1-weighted (T1w) images was performed using ‘N4BiasFieldCorrection’ (Tustison et al., 2010). T1w images were distributed with ANTs 2.3.3 (RRID: SCR_004757; Avants et al., 2008), and used as the T1w-reference throughout the workflow. Skull-stripping of each T1w-reference was applied with a *Nipype* implementation of the ‘antsBrainExtraction.sh’ workflow from ANTs, having OASIS-30ANTs as the target template. On the brain-extracted T1w, brain tissue segmentation of white matter, gray matter, and CSF was done with ‘fast’ (FSL 6.0.5.1:57b01774; RRID: SCR_002823; Zhang et al., 2021). ‘recon-all’ (in OASIS-3: *FreeSurfer* 6.0.1; in ADNI: *FreeSurfer* 7.2.0; RRID: SCR_001847; Dale et al., 1999) was used for brain surface reconstruction. The previously estimated brain mask was refined with a custom variation of the method to reconcile segmentations of the cortical gray matter of Mindboggle derived from ANTs and *FreeSurfer* (RRID: SCR_002438; Klein et al., 2017).

### 2.6 Structural MRI Data

For both data sets, the volume of grey matter regions, including accumbens, amygdala, caudate, hippocampus, pallidum, putamen, and thalamus, were extracted per hemisphere. For all these structures, decreasing volume over the course of AD has previously been observed (Roh et al., 2011; Tang et al., 2014; Yi et al., 2016), albeit often inconsistently. Additionally, structural MRI data included global measurements of total gray matter volume, total subcortical gray matter volume, left and right lateral, 3^rd^, and 4^th^ ventricle volumes, left and right hemisphere mean cortical thickness, left and right total cortical volume, left and right cerebral white matter volume, left and right cerebellar white matter volume, left and right cerebellar cortical volume as well as the volume of five regions of the corpus callosum: anterior, mid-anterior, central, mid-posterior and posterior. Volumetric values were normalized by estimated intracranial volume, given by the volume distortion resulting from the affine registration from the native to the MNI305 standard space, to account for head-size effects. It has previously been shown by Vieira et al. (2022) that this parsimonious feature set performs equally well in predicting cognitive decline as a more extensive set that includes 331 features from global, subcortical, and cortical (volume and thickness) markers.

### 2.7 Treatment of Missing Data

Features with more than 25% missing data in either the ADNI or OASIS-3 dataset were excluded from both cohorts. In total, 18 non-brain features were excluded, reducing the total number from 66 to 48. Removing 10 features pertaining to neuropsychology test items, 4 features recapitulating family history of dementia and 4 features concerning smoking habits, lowered the number of neuropsychological features from 18 to 8 and the number of health information features from 17 to 9. No structural MRI features were excluded (see Supplementary Material, Table B). After removing the features with excessive missing values, 4.0% of non-brain data was missing in OASIS-3 and 4.2% of non-brain data was missing in ADNI prior to imputation.

Imputation is a technique allowing the use of data with missing values. Our predictive pipeline included the IterativeImputer, a multivariate imputer from *Scikit-learn* (Pedregosa et al., 2011). The combination of multiple imputation with predictive modeling has been shown to work in different missingness scenarios (Josse et al., 2019).

### 2.8 Predictive Analysis

The predictive model used in this study was a multi-target random forest (RF) regression model (Breiman, 2001) predicting the rate of decline of CDR-SOB and MMSE simultaneously. The RF algorithm is a non-parametric ML algorithm, which has been shown to excel for tabular data compared to other modern ML alternatives (Grinsztajn et al., 2022; Hengl et al., 2018). It is an ensemble learning method based on decision trees trained on bootstrapped samples (Baez-Villanueva et al., 2020). When employing RF for regression, trees are grown depending on a random vector, hence, the output consists of numerical values instead of class labels as in RF for classification (Breiman, 2001). In this study, we chose to ensemble 100 fully grown trees per model, the default values for RandomForestRegressor from *Scikit-learn* (Pedregosa et al., 2011). All predictive analyses were implemented in *Scikit-learn* 1.0.2 (Pedregosa et al., 2011) in Python 3.9.13.

Predictive models were either trained and tested within the same dataset (*prediction within dataset*) or trained on one dataset and tested on the other (*prediction across datasets*). Three modalities were used for predictions within dataset: non-brain data only (*non-brain model*), structural MRI data only (*structural MRI model*), and the combination of non-brain data and structural MRI data (*combined model*). For predictions across datasets, a fourth modality was introduced: the *top 15 model*. This condition included only the 15 most predictive features of the respective training set, which were previously derived from analyzing the permutation importance of the non-brain features and structural MRI features for predictions within dataset (see Permutation Importance). As ML models can entail biases and may generalize poorly across different subgroups in the population (R. Wang et al., 2023), model performance of the combined model in predictions across datasets was evaluated across subgroups separated by diagnosis, sex, genetic risk (i.e., APOE ε4 allele count) and age.

When training and testing within dataset, data was randomly split into 80% training and 20% test sets using ShuffleSplit. Splitting was repeated 1000 times. For predictions across datasets, the model was trained on the full sample of one dataset and tested on the full sample of the other, meaning that either the OASIS-3 dataset served as the training set and the ADNI dataset as the test set, or vice versa. Full datasets were used to obtain the best possible model. The step of evaluating the prediction capability of the models on independent test data leads to the determination of its generalization performance (Hastie et al., 2009).

### 2.9 Model Evaluation

The models predicting within and across datasets were evaluated primarily using the out-of-sample coefficient of determination (*R^2^*). *R^2^* is often the suggested standard metric for the evaluation of regression analyses. We further evaluated performance with the mean squared error (*MSE*) and the mean absolute error (*MAE*), which are commonly used in ML studies. For predictions within dataset, the evaluation metrics were calculated for each of the 1000 splits. For predictions across datasets, only one test set is available, therefore we measure and compare the distribution of the absolute error (AE), computed for each subject.

### 2.10 Model Comparison

In order to assess whether one model predicted continuous cognitive decline significantly better than another, model comparison was used. As in Vieira et al. (2022), for model comparisons within the same dataset, the number of splits for which the model in question outperformed the reference model, according to *R^2^*, was calculated, resulting in a percentage value. Furthermore, two-sided Wilcoxon signed-rank tests (α = .05) were performed to test whether the differences in model performance were significant (as in, e.g., Bijl-Hofland et al., 1999). A two-sided Mann-Whitney U test (α = .05) was used to test for differences regarding the distributions of *R^2^* between models predicting within OASIS-3 and models predicting within ADNI. To compare the performance of two models that predicted continuous cognitive decline across datasets, the distributions of the *AE* were analyzed with a two-sided Mann-Whitney U test (α = .05). When comparing a model for prediction within dataset to a model for prediction across datasets, in a first step, the differences of *R^2^* were calculated for each split by using subtraction. Next, two-sided Wilcoxon signed-rank tests (α = .05) were performed with the adapted null hypothesis that mean difference is zero (zero_method = “wilcox”). For the evaluation of model performance in different subgroups, two-sided Mann-Whitney U tests (α = .05) for comparison of absolute errors were employed. The *p* values were corrected for multiple comparisons using the Holm-Bonferroni method.

### 2.11 Permutation Importance

Permutation importance was calculated through permuting feature values to evaluate the influence of the features on the predictive performance of prediction within dataset. Thereby, the difference in the predictive performance, as measured by the *R^2^* score, before and after the permutation of the predictive feature is used as a measure of importance. Hence, whenever the shuffling of a feature led to a decrease in performance, the feature was deemed important for the model (Breiman, 2001; Strobl et al., 2008). This procedure of evaluating the importance of input features can increase the interpretability of the model (Woo et al., 2017). For the top 15 model, used only for predictions across datasets, the focus was on the 15 most predictive features of the respective training set. Importantly, the feature selection process was restricted to the training set, ensuring that no information from the test set influenced the selection, thereby avoiding circular analysis. Fifteen features were selected due to the diminishing importance of additional features (Tam et al., 2022; Vieira et al., 2022). Additionally, in a supplementary analysis the permutation importance was computed for predictions across datasets with 100 repetitions.

## 3 Results

### 3.1 Differences Between the Datasets

The sample characteristics of the two datasets are listed in Table 1. There were significant differences regarding sex (*Χ*^2^ = 6.555, *p* = .01), diagnoses (*Χ*^2^ = 440.064, *p* < .001), and education (*t*(1846) = 3.7, *p* < .001) between OASIS-3 and ADNI. In OASIS-3, there were proportionally more women, more HC and less individuals with MCI, and the participants had fewer years of education compared to the ADNI dataset. No significant difference was found concerning age (*t*(1896) = 0.9, *p* = .39). In addition, the expected yearly rate of change in CDR-SOB (*M_OASIS-3_* = 0.25, *M_ADNI_* = 0.46, *t*(1897) = 5.9, *p* < .001) and MMSE (*M_OASIS-3_* = −0.32, *M_ADNI_* = −0.67, *t*(1897) = −6.1, *p* < .001) differed significantly between OASIS-3 and ADNI, with greater rates of decline being observed in ADNI. These differences are illustrated in the Supplementary Material, Figure B, which displays the distribution of CDR-SOB and MMSE change in OASIS-3 and ADNI. Overall, the sample characteristics differed substantially between OASIS-3 and ADNI, illustrating realistically the typical variations observed across independent neuroimaging cohorts, and the importance of validation on independent datasets.

### 3.2 Prediction of Continuous Cognitive Decline Within Dataset

The combined model led to the best prediction of continuous cognitive decline for both CDR-SOB and MMSE in OASIS-3 (CDR-SOB: *R^2^* = .42, *MSE* = .21, *MAE* = .24; MMSE: *R^2^* = .33, *MSE* = .56, *MAE* = .43; see Table 2).^2^ Model comparison showed that the combined model significantly outperformed the non-brain model (CDR-SOB: *R^2^* = .35, *MSE* = .24, *MAE* = .26; MMSE: *R^2^* = .29, *MSE* = .61, *MAE* = .44) in 88.8% of the splits for CDR-SOB, *p <* .001, and in 81.3% of the splits for MMSE, *p <* .001. Further, the combined model significantly outperformed the structural MRI model in 97.2% of the splits for CDR-SOB, *p <* .001, and 96.3% of the splits for MMSE, *p <* .001. Additionally, model comparison between the structural MRI model (CDR-SOB: *R^2^* = .23, *MSE* = .28, *MAE* = .30; MMSE: *R^2^* = .14, *MSE* = .74, *MAE* = .53) and the non-brain model demonstrated that prediction with only structural MRI data performed significantly worse than prediction with only non-brain data in 82.8% of the splits for CDR-SOB, *p <* .001, and in 87.8% for MMSE, *p <* .001. Performance for the prediction of CDR-SOB and MMSE slopes within OASIS-3 are shown in Figure 1.A and Figure 1.B, respectively.

**Figure 1.**
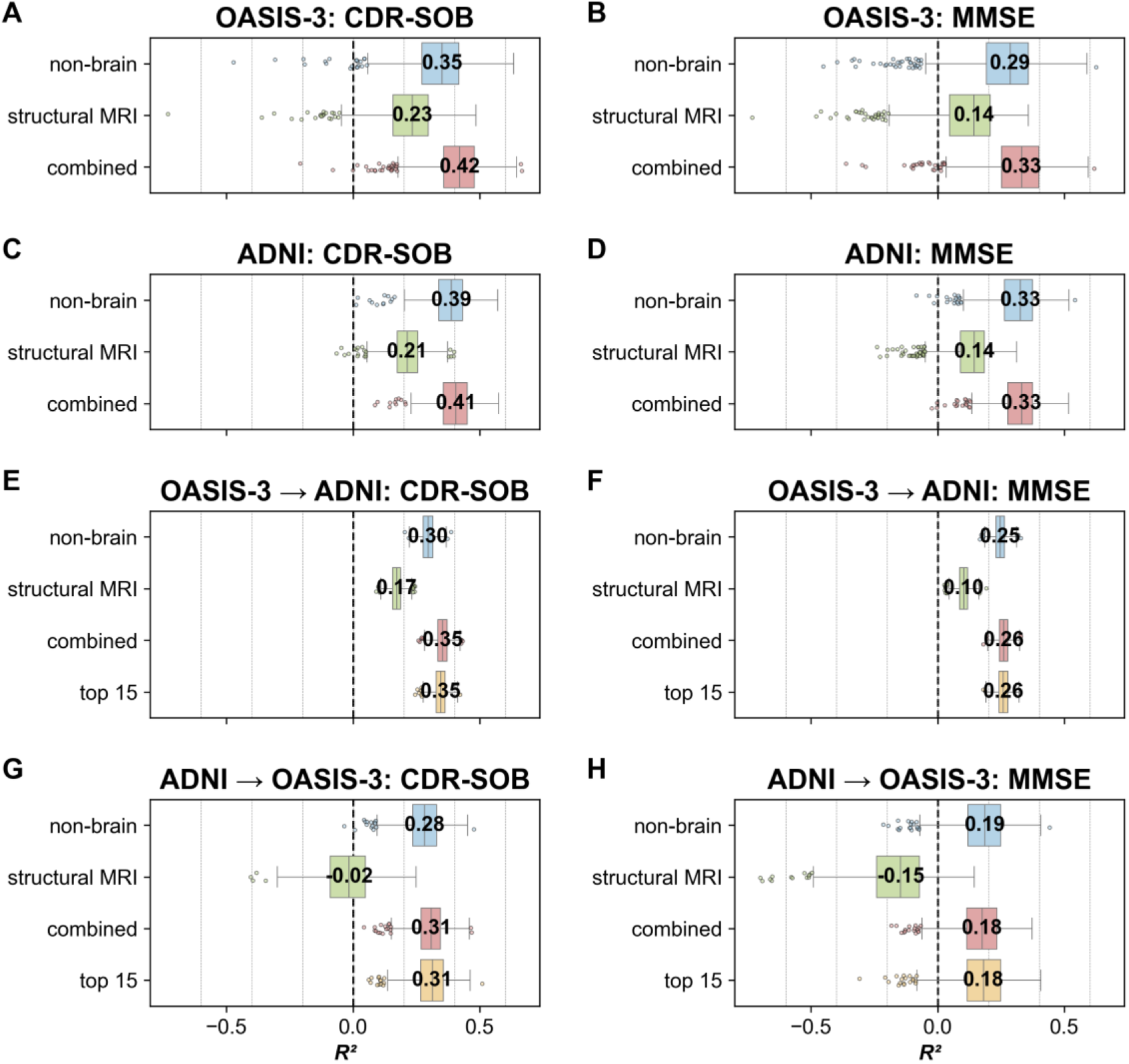
Model Performance in OASIS-3 and ADNI. *Note*. Model performance (*R^2^*, coefficient of determination; x-axis) for the input modalities (non-brain; structural MRI; combined, top 15; y-axis) to predict continuous cognitive decline measured via CDR-SOB and MMSE within and across datasets. For the predictions within dataset (A‒D), *R^2^* is measured across splits (*N*_splits_ = 1000), while for the predictions across datasets (E‒H), *R^2^*is measured across bootstrapped sample (*N*_boot_ = 1000). The number represents the median *R^2^* of the respective model. The dashed vertical line marks *R^2^ =* 0.

**Table 2.** Model Performance and Model Comparisons for Predicting CDR-SOB Change.

| Dataset | Modality | $R^2$ | $MSE$ | $MAE$ | OASIS-3 | | | ADNI | | | OASIS-3 → ADNI | | ADNI → OASIS-3 | |
| --- | --- | --- | --- | --- | --- | --- | --- | --- | --- | --- | --- | --- | --- | --- |
|  |  |  |  |  | 1 | 2 | 3 | 1 | 2 | 3 | 3 | 4 | 3 |  |
| OASIS-3 | 1 non-brain | .35 | .24 | .26 |  |  |  |  |  |  |  |  |  |  |
|  | 2 structural MRI | .23 | .28 | .30 | < .001<sup>a</sup> |  |  |  |  |  |  |  |  |  |
|  | 3 combined | .42 | .21 | .24 | < .001<sup>a</sup> | < .001<sup>a</sup> |  |  |  |  |  |  |  |  |
| ADNI | 1 non-brain | .39 | .57 | .42 | < .001<sup>b</sup> |  |  |  |  |  |  |  |  |  |
|  | 2 structural MRI | .21 | .71 | .52 |  | < .001<sup>b</sup> |  | < .001<sup>a</sup> |  |  |  |  |  |  |
|  | 3 combined | .41 | .55 | .41 |  |  | .001<sup>b</sup> | < .001<sup>a</sup> | < .001<sup>a</sup> |  |  |  |  |  |
| OASIS-3<br>→ ADNI | 1 non-brain | .30 | .65 | .50 | < .001<sup>c</sup> |  |  | < .001<sup>c</sup> |  |  |  |  |  |  |
|  | 2 structural MRI | .17 | .77 | .49 |  | < .001<sup>c</sup> |  |  | < .001<sup>c</sup> |  |  |  |  |  |
|  | 3 combined | .35 | .60 | .45 |  |  | < .001<sup>c</sup> |  |  | < .001<sup>c</sup> |  |  |  |  |
|  | 4 top 15 | .34 | .61 | .42 |  |  |  |  |  |  | < .001<sup>d</sup> |  |  |  |
| ADNI →<br>OASIS-3 | 1 non-brain | .28 | .27 | .29 | < .001<sup>c</sup> |  |  | < .001<sup>c</sup> |  |  |  |  |  |  |
|  | 2 structural MRI | -.02 | .38 | .41 |  | < .001<sup>c</sup> |  |  | < .001<sup>c</sup> |  |  |  |  |  |
|  | 3 combined | .31 | .26 | .30 |  |  | < .001<sup>c</sup> |  |  | < .001<sup>c</sup> | < .001<sup>d</sup> |  |  |  |
|  | 4 top 15 | .31 | .26 | .27 |  |  |  |  |  |  |  | < .001<sup>d</sup> | < .001<sup>d</sup> |  |

*Note.* The model performance and the $p$ values of model comparisons for predicting cognitive decline measured via Clinical Dementia Rating Scale Sum of Boxes (CDR-SOB) are displayed. OASIS-3 → ADNI = model trained on OASIS-3 and tested on ADNI; ADNI → OASIS-3 = model trained on ADNI and tested on OASIS-3. $R^2$ = median coefficient of determination across 1000 splits; $MSE$ = average mean squared error across 1000 splits; $MAE$ = average mean absolute error across 1000 splits.
<sup>a</sup> Two-sided Wilcoxon signed-rank tests ( $\alpha = .05$ ) for comparison of $R^2$ (coefficient of determination). <sup>b</sup> Two-sided Mann-Whitney U test ( $\alpha = .05$ ) for comparison of $R^2$ . <sup>c</sup> Two-sided Wilcoxon signed-rank tests ( $\alpha = .05$ ) with the adapted null hypothesis for comparison of $R^2$ . <sup>d</sup> Two-sided Mann-Whitney U test ( $\alpha = .05$ ) for comparison of absolute errors.

Next, continuous cognitive decline was predicted within the ADNI dataset. In ADNI, adding structural MRI data to non-brain data led to a statistically significant improvement of the prediction for CDR-SOB (*R^2^* = .406, *MSE* = .55, *MAE* = .41, *p* = <.001) and MMSE (*R^2^* = .330, *MSE* = 1.76, *MAE* = .76, *p* = <.001), as compared to the non-brain model (CDR-SOB: *R^2^* = .388, *MSE* = .57, *MAE* = .42; MMSE: *R^2^* = .327, *MSE* = 1.78, *MAE* = .76). The combined model outperformed the non-brain model in 72.3% of the splits for CDR-SOB and 55.6% of the splits for MMSE. Another model comparison revealed that, as in OASIS-3, the combined model showed better model performance than the structural MRI model in 99.8% of the splits for CDR-SOB, *p <* .001, and 99.1% of the splits for MMSE, *p <* .001. Moreover, the structural MRI model (CDR-SOB: *R^2^* = .21, *MSE* = .71, *MAE* = .52; MMSE: *R^2^* = .14, *MSE* = 2.24, *MAE* = .93) performed worse than the non-brain model in 98.6% of the splits for CDR-SOB, *p <* .001, and 97.8% of the splits for MMSE, *p <* .001. Performance for the prediction of CDR-SOB and MMSE slopes within ADNI are shown in Figure 1.C and Figure 1.D, respectively.

Subsequently, the performance of models predicting continuous cognitive decline within the ADNI dataset and within the OASIS-3 dataset was compared by testing for differences regarding the distributions of *R^2^* between models. For histograms of *R^2^* for each split in all models see Supplementary Material, Figures C and D. For the combined model, prediction within OASIS-3 was significantly better than within ADNI for CDR-SOB, *p* = .001, while for MMSE there was no significant difference to be found, *p* = .999. No significant differences in performance were found between ADNI and OASIS-3 when employing only structural MRI data or non-brain data in the prediction of both CDR-SOB and MMSE.

### 3.3 Prediction of Continuous Cognitive Decline Across Datasets

To assess the generalizability of the model across datasets, models were first trained on the OASIS-3 dataset and subsequently evaluated on the independent ADNI dataset. The non-brain model had an *R^2^* = .30, *MSE* = .65, and *MAE* = .50 for CDR-SOB and an *R^2^* = .25, *MSE* = 1.99, and *MAE* = .81 for MMSE. The model performance of the structural MRI model was *R^2^* = .17, *MSE* = .77, and *MAE* = .49 for CDR-SOB and *R^2^* = .10, *MSE* = 2.37, and *MAE* = .89 for MMSE. Furthermore, the combined model demonstrated an *R^2^* = .35, *MSE* = .60, and *MAE* = .45 for CDR-SOB and an *R^2^* = .26 *MSE* = 1.95, and *MAE* = .79 for MMSE.

Likewise, models were then trained on ADNI and evaluated on OASIS-3. For the non-brain model, model performance was *R^2^* = .28, *MSE* = .27, and *MAE* = .29 for CDR-SOB and *R^2^* = .19, *MSE* = .71, and *MAE* = .48 for MMSE. The structural MRI model had an *R^2^* = −.02, *MSE* = 0.38, and *MAE* = .41 for CDR-SOB and an *R^2^* = −.15, *MSE* = 1.00, and *MAE* = .68 for MMSE. Finally, the combined model showed a performance of *R^2^* = .31, *MSE* = .26, and *MAE* = .30 for CDR-SOB and of *R^2^* = 0.18, *MSE* = 0.72, and *MAE* = 0.53 for MMSE. Supplementary Material, Figure E shows the true versus predicted values of CDR-SOB and MMSE change derived from combined models to predict continuous cognitive decline across datasets.

Model comparisons demonstrated that for the three modalities, i.e., non-brain, structural MRI, and combined, models predicting continuous cognitive decline within OASIS-3 were significantly better than models trained on OASIS-3 and tested on ADNI as well as models trained on ADNI and tested on OASIS-3 for both CDR-SOB and MMSE (*p <* .001 for each comparison). Likewise, models predicting continuous cognitive decline within ADNI significantly outperformed models trained on OASIS-3 and tested on ADNI as well as models trained on ADNI and tested on OASIS-3 for both CDR-SOB and MMSE for the three modalities (*p <* .001 for each comparison). The comparison of combined models predicting continuous cognitive decline across datasets showed that the model trained on OASIS-3 and tested on ADNI performed significantly better than the model trained on ADNI and tested on OASIS-3 for CDR-SOB, *p <* .001, and MMSE, *p =* .001. An overview of the model performance of all models and *p* values in model comparisons is provided in Table 2 for predicting CDR-SOB change and Table 3 for predicting MMSE change.

**Table 3.** Model Performance and Model Comparisons for Predicting MMSE Change.

| Dataset | Modality | $R^2$ | MSE | MAE | OASIS-3 | | | ADNI | | | OASIS-3 → ADNI | | ADNI → OASIS-3 |
| --- | --- | --- | --- | --- | --- | --- | --- | --- | --- | --- | --- | --- | --- |
|  |  |  |  |  | 1 | 2 | 3 | 1 | 2 | 3 | 3 | 4 | 3 |
| OASIS-3 | 1 non-brain | .29 | .61 | .44 |  |  |  |  |  |  |  |  |  |
|  | 2 structural MRI | .14 | .74 | .53 | < .001<sup>a</sup> |  |  |  |  |  |  |  |  |
|  | 3 combined | .33 | .56 | .43 | < .001<sup>a</sup> | < .001<sup>a</sup> |  |  |  |  |  |  |  |
| ADNI | 1 non-brain | .33 | 1.78 | .76 | < .001<sup>b</sup> |  |  |  |  |  |  |  |  |
|  | 2 structural MRI | .14 | 2.24 | .93 |  | .201<sup>b</sup> |  | < .001<sup>a</sup> |  |  |  |  |  |
|  | 3 combined | .33 | 1.76 | .76 |  |  | .999<sup>b</sup> | < .001<sup>a</sup> | < .001<sup>a</sup> |  |  |  |  |
| OASIS-3<br>→ ADNI | 1 non-brain | .25 | 1.99 | .81 | < .001<sup>c</sup> |  |  | < .001<sup>c</sup> |  |  |  |  |  |
|  | 2 structural MRI | .10 | 2.37 | .89 |  | < .001<sup>c</sup> |  |  | < .001<sup>c</sup> |  |  |  |  |
|  | 3 combined | .26 | 1.95 | .79 |  |  | < .001<sup>c</sup> |  |  | < .001<sup>c</sup> |  |  |  |
|  | 4 top 15 | .26 | 1.96 | .80 |  |  |  |  |  |  | .233<sup>d</sup> |  |  |
| ADNI →<br>OASIS-3 | 1 non-brain | .19 | .71 | .48 | < .001<sup>c</sup> |  |  | < .001<sup>c</sup> |  |  |  |  |  |
|  | 2 structural MRI | -.15 | 1.00 | .68 |  | < .001<sup>c</sup> |  |  | < .001<sup>c</sup> |  |  |  |  |
|  | 3 combined | .18 | .72 | .53 |  |  | < .001<sup>c</sup> |  |  | < .001<sup>c</sup> | .001<sup>d</sup> |  |  |
|  | 4 top 15 | .18 | .71 | .47 |  |  |  |  |  |  |  | < .001<sup>d</sup> | < .001<sup>d</sup> |

*Note.* The model performance and the $p$ values of model comparisons for predicting cognitive decline measured via Mini-Mental State Examination (MMSE) are displayed. OASIS-3 → ADNI = model trained on OASIS-3 and tested on ADNI; ADNI → OASIS-3 = model trained on ADNI and tested on OASIS-3. $R^2$ = median coefficient of determination across 1000 splits; MSE = average mean squared error across 1000 splits; MAE = average mean absolute error across 1000 splits.
<sup>a</sup> Two-sided Wilcoxon signed-rank tests ( $\alpha = .05$ ) for comparison of $R^2$ (coefficient of determination). <sup>b</sup> Two-sided Mann-Whitney U test ( $\alpha = .05$ ) for comparison of $R^2$ . <sup>c</sup> Two-sided Wilcoxon signed-rank tests ( $\alpha = .05$ ) with the adapted null hypothesis for comparison of $R^2$ . <sup>d</sup> Two-sided Mann-Whitney U test ( $\alpha = .05$ ) for comparison of absolute errors.

### 3.4 Evaluating Model Performance in Different Subgroups

Considering a potential lack of generalizability and common biases in model predictions across subgroups, the combined models’ performance across datasets was assessed within subgroups defined by diagnosis, sex, APOE ε4 status, and age. Statistical testing revealed significant differences between males and females, individuals with MCI or dementia and normal controls, homozygous APOE ε4 carriers and heterozygous carriers or non-carriers, as well as different age groups (see Supplementary Material, Table C). The *MAE* of predicting CDR-SOB and MMSE increases with age, cognitive impairment severity, and APOE ε4 allele dose, and is higher in males than females. Figure 2 shows the *AE* distribution across subgroups.

**Figure 2.**
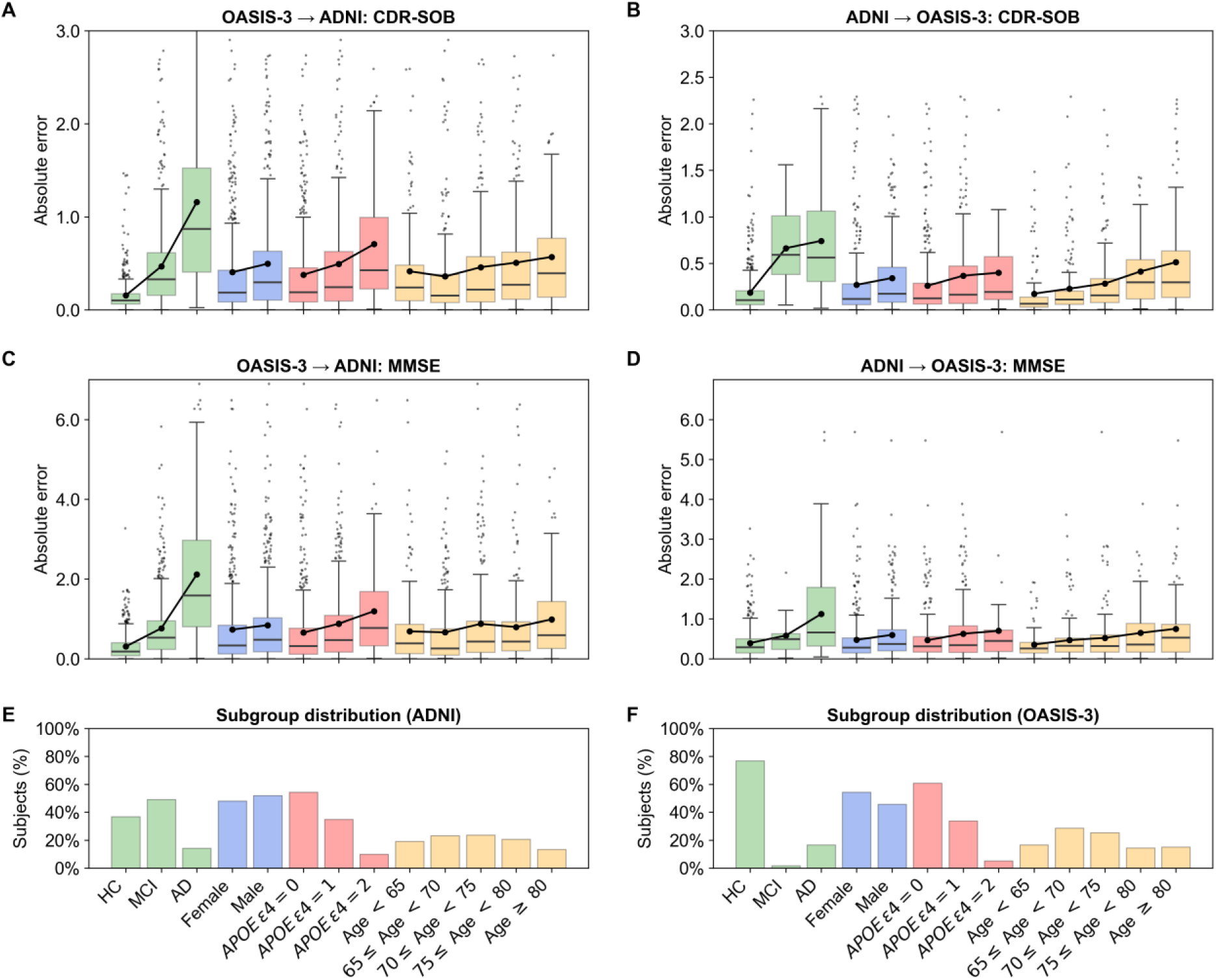
Distribution of Absolute Error Across Subgroups. *Note*. Distribution of absolute error across subgroups in combined models predicting CDR-SOB and MMSE across datasets. Box plots represent the error distributions for CDR-SOB (A, B) and MMSE (C, D) when trained on OASIS-3 and tested on ADNI (A, C) and when trained on ADNI and tested on OASIS-3 (A, D). Dots connected by lines show the average absolute error. Bar plots visualize the distribution of the subgroups in ADNI (E) and OASIS-3 (F).

### 3.5 Most Predictive Features

Permutation importance was used to characterize the most predictive features of the best-performing modality, which was the combined model for both predictions within OASIS-3 and within ADNI. For OASIS-3, the baseline scores of the targets CDR-SOB and MMSE were part of the 15 most important features. Further, the top 15 features of OASIS-3 included neuropsychological scores, such as word fluency and parts of the WMS-R and the TMT, as well as the sum score from the FAQ, which is a clinical feature. From the structural MRI features, the volume of the left and right hippocampus and left and right amygdala along with the central corpus callosum, the volume of the right thalamus and the volume of the right nucleus accumbens were of importance.

In ADNI, the baseline score of the target feature MMSE, but not CDR-SOB, ranks among the 15 most important features. Moreover, neuropsychological scores, such as word fluency, BNT, and parts of the WMS-R and the TMT, together with clinical scores from NPI-Q and FAQ were included among the most important features. Remaining important features in ADNI comprise chronological age at baseline and structural MRI features, namely the volumes of the right hippocampus and 4^th^ ventricle along with the left hemisphere mean cortical thickness. An overview of the top 15 features of OASIS-3 and ADNI is provided in Table 4. In Table D of the Supplementary Material, the permutation importance of all features is listed.

**Table 4.** Most Predictive Features of OASIS-3 and ADNI.

| OASIS-3 |  | ADNI |  |
| --- | --- | --- | --- |
| Feature | PI | Feature | PI |
| MMSE (baseline) | 0.072 (0.069, 0.075) | TMT B TRAIL B | 0.040 (0.038, 0.042) |
| WMS R Logical Memory (MEMUNITS) | 0.071 (0.067, 0.075) | MMSE (baseline) | 0.031 (0.030, 0.032) |
| FAQ (sum score) | 0.053 (0.049, 0.057) | WMS R Logical Memory (MEMUNITS) | 0.030 (0.028, 0.031) |
| *Hippocampus volume (left)* | 0.027 (0.026, 0.029) | *FAQ (bills)* | 0.013 (0.012, 0.014) |
| WMS R Logical Memory (LOGIMEM) | 0.018 (0.016, 0.019) | WMS R Logical Memory (LOGIMEM) | 0.012 (0.011, 0.013) |
| TMT B TRAIL B | 0.013 (0.012, 0.014) | *TMT A TRAIL A* | 0.0090 (0.0075, 0.011) |
| *CDR-SOB (baseline)* | 0.0068 (0.0050, 0.0086) | FAQ (sum score) | 0.0075 (0.0061, 0.0088) |
| *Amygdala volume (left)* | 0.0057 (0.0043, 0.0070) | *BNT* | 0.0062 (0.0053, 0.0071) |
| Word fluency (animals) | 0.0050 (0.0041, 0.0058) | Word fluency (animals) | 0.0061 (0.0043, 0.0079) |
| *Nucleus accumbens volume (right)* | 0.0045 (0.0039, 0.0051) | *Mean cortical thickness (left)* | 0.0061 (0.0055, 0.0066) |
| *Central portion of the Corpus Callosum volume* | 0.0043 (0.0035, 0.0050) | *NPI-Q (severity)* | 0.0048 (0.0042, 0.0054) |
| *Amygdala volume (right)* | 0.0040 (0.0032, 0.0049) | *Age at baseline* | 0.0039 (0.0035, 0.0044) |
| *TMT B TRAIL B / TRAIL A* | 0.0037 (0.0032, 0.0042) | Hippocampus volume (right) | 0.0037 (0.0034, 0.0041) |
| Hippocampus volume (right) | 0.0037 (0.0029, 0.0044) | *4<sup>th</sup> ventricle volume* | 0.0036 (0.0032, 0.0040) |
| *Thalamus volume (right)* | 0.0032 (0.0026, 0.0039) | *FAQ (taxes)* | 0.0030 (0.0024, 0.0037) |

Note. Permutation importance of the top 15 features of the combined models for prediction within dataset, ordered by importance. Features unique to either ADNI or OASIS-3 are shown in italics. PI = mean permutation importance across 1000 splits.

Additionally, Figure G of the Supplementary Material shows the feature importance for predictions across datasets. Ten features deemed important for predictions within OASIS-3 also emerged as important when the model trained on OASIS-3 data was applied to ADNI data. Similarly, 8 out of the 15 most important features for predictions within ADNI overlapped with those identified when the model trained on ADNI was applied to OASIS-3 data.

### 3.6 Prediction of Cognitive Decline with Top 15 Model Across Datasets

Given the potentially substantial information overlap among the original features, models using only the top 15 features from each training set were evaluated to test whether more parsimonious models could maintain predictive performance across datasets. New models including only the top 15 features of the respective training set were used for the prediction of continuous cognitive decline across datasets. To avoid circular analysis, we did not perform predictions within dataset using the top 15 models.

The model trained on the top 15 features of OASIS-3 and tested on the ADNI dataset resulted in a model performance of *R^2^* = .34, *MSE* = .61, and *MAE* = .42 for CDR-SOB and *R^2^* = .26, *MSE* = 1.96, and *MAE* = .80 for MMSE. The model trained on the top 15 features of ADNI and tested on the OASIS-3 dataset achieved an *R^2^* = .31, *MSE* = .26, and *MAE* = .27 for CDR-SOB and *R^2^* = .18, *MSE* = .71, and *MAE* = .47 for MMSE.

Note that the importance of the 15^th^ feature, especially in ADNI, approached zero.

Next, for predictions across datasets, top 15 models were compared to combined models by analyzing the difference in distributions of the *AE*. Model comparison showed that for models trained on OASIS-3 and tested on ADNI the top 15 model outperformed the combined model for CDR-SOB (*p* <.001), whereas for MMSE, there was no significant difference in performance (*p* = .23). When models were trained on ADNI and tested on OASIS-3, the top 15 model performed significantly better than the combined model for both CDR-SOB (*p* <.001) and MMSE (*p <* .001). Tables 2 and 3 provide an overview of the model performance of all models and display the *p* values of the model comparisons for models predicting CDR-SOB and MMSE change, respectively.

## 4 Discussion

This study investigated whether adding structural MRI data to non-brain data significantly improved the performance of an ML model predicting continuous cognitive decline and analyzed the models’ generalizability within and across datasets. Our findings revealed that in ADNI, adding structural MRI data to non-brain data significantly improved the prediction within dataset for both MMSE and CDR-SOB, although modestly for the latter. Further, the performance of models for prediction within dataset differed significantly between ADNI and OASIS-3. Models predicting continuous cognitive decline within one dataset (OASIS-3 or ADNI) performed significantly and substantially better than models predicting continuous cognitive decline across datasets in any modality (see Figure 1 and Tables 2 and 3).

Evaluating the potential of structural MRI data to enhance performance in the prediction of continuous cognitive decline, the current study demonstrated that adding structural MRI data to non-brain data significantly improved the prediction of continuous cognitive decline when assessed with CDR-SOB or MMSE in the ADNI dataset. The improvement in prediction through the addition of structural MRI data is in line with previous studies predicting cognitive decline categorically (Visser et al., 2002) and continuously (Vieira et al., 2022) as well as predicting conversion from MCI to AD (Korolev et al., 2016) and membership in data-driven trajectory-groups of future decline (Bhagwat et al., 2018; Tam et al., 2022).

To reduce the gap between research and the real world, assessing between-site and within-site generalizability plays an important role (Cai et al., 2020; Yarkoni & Westfall, 2017). Our results showed that model performance was inferior for predictions across datasets than for predictions within datasets, as expected. Thus, important considerations arise when applying said models to new, unseen cohorts (Myszczynska et al., 2020; Woo et al., 2017). This is a common finding in machine-learning as some of the statistical patterns relating input features to outputs learned within one dataset might be idiosyncratic to it, with no correspondence in a new dataset. No matter how good one’s model seems to be for prediction within dataset, the performance estimates in the original dataset are likely optimistic (Yarkoni & Westfall, 2017).

The minimization of overfitting is deemed one of the main problems in ML (Domingos, 2012). Having a larger sample size is regarded as a protective factor against overfitting. The datasets used in this study, consisting of 662 participants in OASIS-3 and 1237 participants in ADNI, can be considered as large. However, there are several factors in this study contributing to an increase in overfitting. For one thing, an increased number of predictors might aggravate overfitting (Yarkoni & Westfall, 2017). In the present case, there were 83 input variables: 48 non-brain features and 35 structural MRI features. Moreover, variability driven by site effects can impact the generalizability of results and lead to bias. For instance, for the retrieval of structural MRI data, there were differences regarding the scanner vendors, models, acquisition parameters, and voxel sizes (see Supplementary Material, Table A) – factors that have been repeatedly shown to affect neuroimaging measurements (Chen et al., 2022). Additionally, there were significant differences in the datasets, for example, concerning the participants’ sex, diagnoses, and education, limiting the models’ generalizability. Therefore, the model performances for predictions across datasets were compared for different subgroups. It was shown that the combined model performed significantly better for HC than for individuals with MCI and AD of the respective test set (see Supplementary Material, Table C). A possible explanation for this could point to the floor and ceiling effects observed in CDR-SOB and MMSE, respectively. For CDR-SOB, strong floor effects were observed, with participants having a baseline score of 0 and no change over time amounting to 55.1% in OASIS-3 and 20.5% in ADNI. Notably, the vast majority of these participants were HC: 99.7% in OASIS-3 and 100% in ADNI. This suggests that the floor effects occurred largely due to HC, for whom cognitive decline is minimal or absent, making their prediction easier for the models. For MMSE, ceiling effects were less pronounced, as participants with a baseline score of 30 and no change over time totaled 6.9% in OASIS-3 and 4.0% in ADNI. Among these participants, 93.5% in OASIS-3 and 86% in ADNI were HC. While ceiling effects in MMSE were less impactful than the floor effects in CDR-SOB, they may still have contributed to the better model performance for HC in the test set. Along with this, it was found that for models predicting cognitive decline across datasets, the predictive performance was significantly better for participants carrying zero APOE ε4 alleles than for participants carrying either one or two APOE ε4 alleles (see Figure 2; Supplementary Material, Table C). This could be a compound effect, since carrying at least one APOE ε4 allele is one of the strongest risk factors for sporadic AD (Scheltens et al., 2021). Performance was also found to decrease with increasing age.

In the current study, the difference regarding the clinical diagnoses of participants in OASIS-3 and ADNI further manifests as a significant difference in the rate of decline of both CDR-SOB and MMSE, with greater slopes in ADNI. As a first step to address this issue, another post hoc analysis was performed. In the post hoc analysis, the distribution of the predictive targets (CDR-SOB and MMSE slope) of the test set was matched to that of the training set in the prediction across datasets. The results showed that this matching procedure led to an improvement in terms of *AE* but an observable reduction in *R^2^* in all the comparisons of models trained on OASIS-3 and tested on ADNI. For models trained on ADNI and tested on OASIS-3, the matching procedure led to worsening concerning *AE* but mostly observable improvements in terms of *R^2^* (see Supplementary Material, Table E). A potential reason for that is that the model trained on OASIS-3 might be more accurate for participants with less severe decline and, conversely, the model trained on ADNI performs better in participants with more severe impairment. Matching the test set to the training set distribution reduces the variance of the outputs in the case of the model trained on OASIS-3 and tested on ADNI and increases the proportion of subjects with some level of impairment, which are usually linked to worse AE, in the case of the model trained on ADNI and tested on OASIS-3. Thus, we demonstrated that performance differences cannot be solely attributed to different distributions of target variables. However, it cannot be ruled out that subsamples built based on matching other variables, such as sex, age, or ethnicity would yield different results, as bias has been demonstrated to be a challenge in this regard (Gao & Cui, 2020; Larrazabal et al., 2020; Li et al., 2022; E. Petersen et al., 2022; R. Wang et al., 2023).

When assessing the predictive value of a restricted model trained on the OASIS-3 dataset predicting continuous cognitive decline in the ADNI dataset, it was shown that the top 15 model performed significantly better than the combined model for CDR-SOB, while there was no significant difference in performance for MMSE. When trained on ADNI and tested on OASIS-3, the top 15 model significantly outperformed the combined model for both CDR-SOB and MMSE. This sheds light on the trade-off between simpler models trained on more parsimonious data compared to more complex models trained on all the available variables, and how these simpler models mitigate the issues of generalizability. Although the composition of the top 15 features varied between OASIS-3 and ADNI, there was an overlap regarding the baseline score of MMSE, the sum score from the FAQ, word fluency, parts of the WMS-R and the TMT, as well as the volume of the right hippocampus. While it is somewhat evident for the baseline score of MMSE to be of importance, as it is included in the calculation of MMSE slope (see Modeling the Prediction of Continuous Cognitive Decline), past work has shown that participants with lower MMSE scores are more likely to progress further towards dementia (Edwin et al., 2021). The baseline CDR-SOB score, however, was identified as one of the most important features of OASIS-3, but not of ADNI. One potential reason for this is that the permutation approach may underestimate the importance of correlated features. This could be explained by the shared information one feature still carries when the correlated feature is permuted (Vieira et al., 2022). The strong negative correlation between CDR-SOB and MMSE (i.e., in OASIS-3: *r*(660) = −0.64; in ADNI: *r*(1235) = −0.72), indicating that higher scores on the CDR-SOB are related to lower scores on the MMSE, and vice versa (Balsis et al., 2015; Linz et al., 2017), could thus justify the baseline CDR-SOB score not falling within the top 15 features of ADNI. A potential solution is to compute feature importance of groups of related variables (Chamma et al., 2023).

Both the CDR-SOB and the MMSE are often used instruments for assessing the severity of AD (Mitchell, 2013; O’Bryant et al., 2008). The CDR was reported to be a useful screening for MCI and dementia, showing high interrater reliability as well as high sensitivity and specificity (Chaves et al., 2007; Huang et al., 2021; Juva et al., 1995). The CDR-SOB, which is obtained by summing each of the CDR box scores enables researchers to monitor the progression of dementia within stages, in addition to between stages, and thus is more precise in tracking changes over time compared to the global CDR score (O’Bryant et al., 2008). Compared to the CDR-SOB, the MMSE is less accurate in pre-screening the severity of cognitive dysfunction (Balsis et al., 2015), but it is seen as a practical method, as it, for instance, does not require a close informant and represents the most used rapid cognitive screening instrument (Juva et al., 1995; Mitchell, 2013). Nonetheless, a limiting aspect of both scores is their floor and ceiling effects elaborated earlier in this discussion (Balsis et al., 2015; Franco-Marina et al., 2010; Linz et al., 2017).

Regarding the other top 15 features, previous research has also revealed associations to the prediction of continuous cognitive decline. FAQ scores were shown to be useful for distinguishing between MCI and AD (Teng et al., 2010) as well as predicting progression from MCI to AD in ADNI data (Berezuk et al., 2023). Word fluency, specifically category fluency, helped to differentiate between MCI and AD (Fernaeus et al., 2008) and predicted the risk of incident MCI (Jacobs et al., 2021). Moreover, WMS-R subtests were presented as predictors for distinguishing individuals with dementia from healthy subjects (Derrer et al., 2001). The TMT was demonstrated to be a predictor of conversion from MCI to dementia (Eckerström et al., 2015; Ewers et al., 2012) and, in line with this, dementia progression (Edwin et al., 2021). Lastly, besides the hippocampal volume being a core AD biomarker (Jack et al., 2011; Pini et al., 2016), the right hippocampus volume is a predictor of conversion from MCI to AD (Herukka et al., 2008; Nesteruk et al., 2015).

### 4.1 Limitations and Future Research

Examining cognitive decline as a continuous variable instead of coercing it into distinct categorical labels can help circumvent the problem of the artificiality and imprecision of dividing the cognitive continuum into healthy aging, MCI and AD (Jack et al., 2010; Plassman et al., 2010). Predicting continuous cognitive decline could enable explanations on an individual level which might be useful for the identification of features relevant to specific disease stages or phenotypes and could aid the development of preventative therapies (Martin et al., 2023). However, for the estimation of cognitive decline as a continuous variable, longitudinal data from various clinical assessments is necessary. In this study, participant-specific linear models were applied to estimate the target variables, but we cannot rule out that non-linear trajectories of future CDR-SOB and MMSE scores might provide a better representation of cognitive decline (Vieira et al., 2022; Wilkosz et al., 2010). Linear trajectories provide a useful approximation of cognitive decline due to being robustly estimated with only three measurements and approximate the immediate rate of change. In contrast, non-linear models are more complex, requiring more longitudinal clinical measurements per participant and while likely less biased are more prone to variance. Therefore, we consider linear models to be a good approximation. Nevertheless, future studies with larger datasets and more longitudinal clinical measurements could investigate the prediction of non-linear trajectories. In addition, while we included imaging data from a single timepoint into a predictive model, aggregating information from multiple longitudinal imaging measurements is worth considering (Marinescu et al., 2021).

ML algorithms, such as RF used in this study, are often deemed as “black boxes”. There is a lack of transparency that hinders the understanding of the addressed issue and potentially reduces the usefulness of the outputs (Myszczynska et al., 2020). To counteract this problem, the importance of input features was evaluated comparing the *R^2^* scores before and after permuting each feature (Strobl et al., 2008; Woo et al., 2017). *R^2^* was chosen as the primary performance metric in this work due to its prominence in the literature and its scale invariance, but the observed findings may not necessarily generalize to other performance metrics that were not assessed in this study. Further, it might be possible that our findings do not generalize to other ML models using different algorithms, for example, minimal recurrent neural networks (Nguyen et al., 2020), which should be investigated by future research (Grinsztajn et al., 2022).

The results regarding the generalizability of the top 15 models demonstrated conserved performance compared to the combined model, based on 83 features. Therefore, future research should consider testing how well such models with a reduced number of features generalize to independent datasets. Deriving from the findings of this study, a model of interest might include the features that showed comparably high predictive value in both OASIS-3 and ADNI, namely the baseline score of MMSE, the sum score from the FAQ, word fluency, parts of the WMS-R and the TMT, as well as the volume of the right hippocampus.

Different patterns of missing data across subjects can affect the identification of important features and blur generalizability estimates. To counteract this issue, we opted to remove 18 non-brain features with an excessive number of missing values. It is still possible that missing value patterns in the remaining features are systematically related to outcomes.

The potential bias entailed in ML models, questioning their impact on clinical settings, is a further limiting factor of this study. ML models have previously been shown to predict differently for different ages (R. Wang et al., 2023), sexes (Larrazabal et al., 2020; E. Petersen et al., 2022; R. Wang et al., 2023), and ethnic groups (Gao & Cui, 2020; Li et al., 2022). To shed light on this issue, this study investigated the performance of the combined model in subsamples differing in terms of diagnosis, sex, APOE ε4 and age, revealing significant performance differences for the distinct subgroups. Even though, using multi-centric large-scale data as well as testing the models on a completely independent sample, both done in the present study, have been shown to mitigate bias, in some circumstances, bias cannot be removed and thus requires further investigation (R. Wang et al., 2023; Woo et al., 2017). Moreover, future research should investigate the impact of matching procedures on model performance across different datasets, particularly focusing on the effects of matching not just predictive targets (as it was done in a post hoc analysis within this study), but also other variables such as sex, age, and ethnicity. Given that the current findings suggest that matching may improve or worsen model performance depending on the dataset and the target variables, it is essential to explore whether matching on additional variables could mitigate potential biases and enhance model generalization. This line of inquiry is crucial for developing robust models that perform consistently across diverse populations and conditions.

### 4.2 Conclusion

This study showed an improvement in the prediction of future continuous cognitive decline when adding structural MRI data to non-brain data in the ADNI cohort, as previously demonstrated for OASIS-3. Further, the performance of models being trained on only non-brain data, only structural MRI data, their combination, or the 15 most predictive features of the training set was tested in an independent dataset. While we attested significant performance reduction in all four conditions, substantial performance degradation was most present in the models trained solely on structural MRI data. Models trained only on the top 15 features of the respective training set obtained virtually the same level of performance as the models trained on all non-brain features and structural MRI features when tested externally. Future studies could therefore incorporate structural MRI data in their predictions and evaluate the performance of a model with a reduced set of important features on an independent dataset. In this study, the performance degradation in across dataset predictions could not be circumvented by matching the distributions of the target variables of the test set to those of the respective training set. Future studies should explore other potential bias mitigation strategies, such as feature harmonization and transfer learning. These steps could be important in developing a generalizable model and thus towards application to cognitive impairment or otherwise in the real world (Myszczynska et al., 2020; Yarkoni & Westfall, 2017). Moreover, the present investigation of cognitive decline as a continuous variable enables more precise predictions on an individual level and contributes to developments in the direction of precision medicine (Martin et al., 2023; Vieira et al., 2022).

## Ethics statement

This study was conducted using publicly available, de-identified data from the OASIS-3 and ADNI cohorts. All participants in both studies provided informed consent, and data collection was approved by the respective local institutional review boards. The use of these datasets complies with their data use agreements and ethical guidelines.

## Data and Code Availability

Part of the data used in preparation for this article was obtained from the Alzheimer’s Disease Neuroimaging Initiative (ADNI) database (adni.loni.usc.edu). To access the publicly and freely available ADNI data from the Laboratory of Neuro Imaging (LONI) Image and Data Archive (IDA) at https://adni.loni.usc.edu/data-samples/adni-data/, an application form including the investigator’s institutional affiliation and the proposed uses of the ADNI data must be submitted.

OASIS-3 is an open-access longitudinal multimodal neuroimaging, clinical, and cognitive dataset for normal aging and Alzheimer’s disease from the Open Access Series of Imaging Studies (OASIS). The OASIS-3 dataset can be accessed at https://www.oasis-brains.org.

The code is available on GitHub and on OSF.

## Author Contributions

All authors conceptualized the study. B. H. V. preprocessed the MRI data. R. M. H. and B. H. V. conducted the machine learning analysis and provided visualization of the data. R. M. H. interpreted the results and wrote the original draft. R. M. H. and B. H. V. revised the manuscript. B. H. V. and N. L. supervised R. M. H. during the whole process. All authors provided critical feedback on the manuscript.

## Data Availability

Part of the data used in preparation for this article was obtained from the Alzheimer's Disease Neuroimaging Initiative (ADNI) database (adni.loni.usc.edu). To access the publicly and freely available ADNI data from the Laboratory of Neuro Imaging (LONI) Image and Data Archive (IDA) at https://adni.loni.usc.edu/data-samples/adni-data/, an application form including the investigator's institutional affiliation and the proposed uses of the ADNI data must be submitted.
OASIS-3, the second dataset that was used, is an open-access longitudinal multimodal neuroimaging, clinical, and cognitive dataset for normal aging and Alzheimer's disease from the Open Access Series of Imaging Studies (OASIS). The OASIS-3 dataset can be accessed at https://www.oasis-brains.org.

https://www.oasis-brains.org/

https://github.com/rhuepp/gemacode.git

https://osf.io/up65f/?view_only=23ec3db71e814d048bb8461b24ad2f87

## Acknowledgements

We want to acknowledge the data provided by the Open Access Series of Imaging Studies (OASIS-3: Longitudinal Multimodal Neuroimaging: Principal Investigators: T. Benzinger, D. Marcus, J. Morris; NIH P30 AG066444, P50AG00561, P30 NS09857781, P01 AG026276, P01 AG003991, R01 AG043434, UL1TR000448, R01 EB009352) as well as the data obtained from the Alzheimer’s Disease Neuroimaging Initiative (ADNI) database (adni.loni.usc.edu).

Data collection and sharing for this project was funded by ADNI (National Institutes of Health Grant U19 AG024904) and DOD ADNI (Department of Defense award number W81XWH-12-2-0012). ADNI is funded by the National Institute on Aging, the National Institute of Biomedical Imaging and Bioengineering, and through generous contributions from the following: AbbVie, Alzheimer’s Association; Alzheimer’s Drug Discovery Foundation; Araclon Biotech; BioClinica, Inc.; Biogen; Bristol-Myers Squibb Company; CereSpir, Inc.; Cogstate; Eisai Inc.; Elan Pharmaceuticals, Inc.; Eli Lilly and Company; EuroImmun; F. Hoffmann-La Roche Ltd and its affiliated company Genentech, Inc.; Fujirebio; GE Healthcare; IXICO Ltd.; Janssen Alzheimer Immunotherapy Research & Development, LLC.; Johnson & Johnson Pharmaceutical Research & Development LLC.; Lumosity; Lundbeck; Merck & Co., Inc.; Meso Scale Diagnostics, LLC.; NeuroRx Research; Neurotrack Technologies; Novartis Pharmaceuticals Corporation; Pfizer Inc.; Piramal Imaging; Servier; Takeda Pharmaceutical Company; and Transition Therapeutics. The Canadian Institutes of Health Research is providing funds to support ADNI clinical sites in Canada. Private sector contributions are facilitated by the Foundation for the National Institutes of Health (www.fnih.org). The grantee organization is the Northern California Institute for Research and Education, and the study is coordinated by the Alzheimer’s Therapeutic Research Institute at the University of Southern California. ADNI data are disseminated by the Laboratory for Neuro Imaging at the University of Southern California. Investigators within the ADNI contributed to the design and implementation of ADNI and/or provided data but did not participate in analysis or writing of this report. A complete listing of ADNI investigators can be found at: http://adni.loni.usc.edu/wp-content/uploads/how_to_apply/ADNI_Acknowledgement_List.pdf.

## Supplementary Material

Supplementary material is available on OSF.

## Footnotes

1 Data were provided by OASIS-3: Longitudinal Multimodal Neuroimaging (Principal Investigators: T. Benzinger, D. Marcus, J. Morris; NIH P30 AG066444, P50 AG00561, P30 NS09857781, P01 AG026276, P01 AG003991, R01 AG043434, UL1 TR000448, R01 EB009352).

2 Please note that all *R^2^* values being described regarding predictions within datasets refer to the median *R^2^* across 1000 splits. Output Tables including the *R^2^*, *MSE*, and *MAE* values for each of the 1000 splits are available on OSF.

