## Supplementary Material for "Predicting Continuous Cognitive Decline: The Generalizability of a Multimodal Machine Learning Approach Including Structural MRI and Non-Brain Data"

**Roya Melanie Hüppi[1, 2, 3], Nicolas Langer[1, 2], Bruno Hebling Vieira[1, 2, ✉] for the Alzheimer's Disease Neuroimaging Initiative***

[1] Methods of Plasticity Research, Department of Psychology, University of Zurich, Zurich, Switzerland

[2] Neuroscience Center Zurich (ZNZ), University of Zurich & ETH Zurich, Zurich, Switzerland

[3] Department of Adult Psychiatry and Psychotherapy, Psychiatric University Clinic Zurich and University of Zurich, Zurich, Switzerland

*Data used in preparation of this article were obtained from the Alzheimer's Disease Neuroimaging Initiative (ADNI) database (adni.loni.usc.edu). As such, the investigators within the ADNI contributed to the design and implementation of ADNI and/or provided data but did not participate in analysis or writing of this report. A complete listing of ADNI investigators can be found at: http://adni.loni.usc.edu/wp-content/uploads/how_to_apply/ADNI_Acknowledgement_List.pdf

### Supplementary Material

Since listing the $R^2$, *MSE*, and *MAE* values for each of the 1000 splits of the models predicting continuous cognitive decline within datasets and all the features' permutation importance would have taken up a lot of space, these values are not listed in the supplementary material. Instead, the output tables are available on OSF.

#### A. Supplementary Tables

#### Table A

*Details on MRI Acquisition in OASIS-3 and ADNI.*

| Dataset<br>Scanner | Voxel-size<br>(mm × mm × mm) | TR (ms) | TE (ms) | Subjects |
|---|---|---|---|---|
| OASIS-3 | | | | |
| Biograph mMR | 1 × 1 × 1 | 2400 | 2.1 | 2 |
| | 1 × 1.05 × 1.05 | 2300 | 3.0 | 42 |
| TrioTim | 1 × 1 × 1 | 2400 | 3.1 | 30 |
| | 1 × 1.02 × 1.02 | 2400 | 3.1 | 1 |
| | 1 × 1 × 1 | 2400 | 3.2 | 590 |
| Missing | 1 × 1 × 1 | | | 1 |
| ADNI | | | | |
| Achieva | 1 × 1 × 1 | | 2.9 | 24 |
| | 1.2 × 1 × 1 | | 3.1 | 120 |
| | 1.2 × 1 × 1 | | 3.2 | 19 |
| | 1.2 × 1 × 1 | | 3.3 | 13 |
| Achieva dStream | 1 × 1 × 1 | | 2.9 | 44 |
| Allegra | 1.2 × 1 × 1 | 2300 | 2.9 | 29 |
| Biograph_mMR | 1 × 1 × 1 | 2300 | 3 | 6 |
| DISCOVERY MR750 | 1 × 1 × 1 | | 3.1 | 62 |
| | 1 × 1 × 1 | | 3.2 | 1 |
| | 1.2 × 1.02 × 1.02 | | 3 | 97 |
| | 1.2 × 1.02 × 1.02 | | 3.2 | 11 |
| | 1.2 × 1.05 × 1.05 | | 3 | 48 |
| DISCOVERY MR750w | 1 × 1 × 1 | | 3.1 | 29 |
| | 1.2 × 1.02 × 1.02 | | 3.1 | 10 |
| | 1.2 × 1.05 × 1.05 | | 3.1 | 1 |
| GEMINI | 1.2 × 1 × 1 | | 3.1 | 15 |
| GENESIS_SIGNA | 1.2 × 1.02 × 1.02 | | 3.1 | 6 |
| Ingenia | 1 × 1 × 1 | | 2.9 | 51 |
| | 1 × 1 × 1 | | 3 | 2 |
| | 1.2 × 1 × 1 | | 3.2 | 18 |
| | 1.2 × 1.05 × 1.05 | | 3.1 | 1 |

**Table A (continued)**

| Dataset<br>Scanner | Voxel-size<br>(mm × mm × mm) | TR (ms) | TE (ms) | Subjects |
|---|---|---|---|---|
| Ingenuity | 1.2 × 1 × 1 | | 3.1 | 3 |
| Intera | 1 × 1 × 1 | | 2.9 | 2 |
| | 1.2 × 1 × 1 | | 3.1 | 13 |
| | 1.2 × 1 × 1 | | 3.2 | 52 |
| | 1.2 × 1 × 1 | | 3.3 | 7 |
| Intera Achieva | 1.2 × 1 × 1 | | 3.3 | 1 |
| Prisma | 1 × 1 × 1 | 2300 | 3 | 77 |
| | 1.2 × 1.05 × 1.05 | 2300 | 3 | 9 |
| Prisma_fit | 1 × 1 × 1 | 2300 | 3 | 195 |
| | 1.2 × 1 × 1 | 2300 | 3 | 1 |
| | 1.2 × 1.05 × 1.05 | 2300 | 3 | 30 |
| | 1.2 × 1.25 × 1.25 | 1800 | 2.5 | 1 |
| SIGNA EXCITE | 1.2 × 1.02 × 1.02 | | 2.8 | 8 |
| | 1.2 × 1.02 × 1.02 | | 3 | 12 |
| SIGNA HDx | 1.2 × 1.02 × 1.02 | | 2.8 | 13 |
| SIGNA Premier | 1 × 1 × 1 | | 2.9 | 8 |
| | 1 × 1 × 1 | | 3 | 4 |
| | 1.3 × 1.05 × 1.05 | | 2.9 | 1 |
| SIGNA UHP | 1 × 1 × 1 | | 3 | 2 |
| Signa HDxt | 1.2 × 1.02 × 1.02 | | 2.8 | 109 |
| | 1.2 × 1.02 × 1.02 | | 3 | 8 |
| | 1.2 × 1.05 × 1.05 | | 2.8 | 8 |
| | 1.2 × 1.05 × 1.05 | | 3 | 2 |
| Skyra | 1 × 1 × 1 | 2300 | 3 | 41 |
| | 1.2 × 1 × 1 | 2300 | 3 | 76 |
| | 1.2 × 1.02 × 1.02 | 2300 | 3 | 1 |
| | 1.2 × 1.05 × 1.05 | 2300 | 3 | 32 |
| Skyra_fit | 1 × 1 × 1 | 2300 | 3 | 2 |
| Trio | 1.2 × 1 × 1 | 2300 | 2.9 | 60 |
| | 1.2 × 1 × 1 | 2300 | 3 | 1 |
| TrioTim | 1 × 1 × 1 | 2300 | 3 | 41 |
| | 1.2 × 1 × 1 | 2300 | 2.9 | 19 |
| | 1.2 × 1 × 1 | 2300 | 3 | 335 |
| | 1.2 × 1.02 × 1.02 | 2300 | 2.9 | 1 |
| | 1.2 × 1.02 × 1.02 | 2300 | 3 | 1 |
| | 1.2 × 1.05 × 1.05 | 2300 | 3 | 3 |
| Verio | 1 × 1 × 1 | 2300 | 3 | 58 |
| | 1.2 × 0.98 × 0.98 | 2300 | 3 | 1 |
| | 1.2 × 1 × 1 | 2000 | 3 | 1 |
| | 1.2 × 1 × 1 | 2300 | 3 | 151 |
| | 1.2 × 1.05 × 1.05 | 2300 | 3 | 2 |
| | 1.2 × 1.09 × 1.09 | 2300 | 2.9 | 2 |

*Note*. Details on MRI acquisition from the OASIS-3 dataset are based on Vieira et al. (2022). Empty cells mean that data is missing. TR = repetition time; TE = echo time.

**Table B**

*Overview of All the Included Features.*

| Modality (# features) | Group (# features) | Test/Variables | Information |
|---|---|---|---|
| Non-brain (48) | Demographic information (3) | Age | |
| | | Sex | |
| | | Education | |
| | Clinical scores (23) | Mini-Mental State Examination (MMSE) | Sum score |
| | | Clinical Dementia Rating Scale Sum of Boxes (CDR-SOB) | 6 items; Global score; SOB score |
| | | Functional Activities Questionnaire (FAQ) | 10 items; Sum score |
| | | Neuropsychiatric Inventory Questionnaire (NPI-Q) | Symptom presence; Severity sum scores |
| | | Geriatric Depression Scale (GDS) | Sum score |
| | Neuropsychological scores (8) | Wechsler Memory Scale-Revised (WMS-R) | Logical memory (LOGIMEM, MEMUNITS, MEMTIME); |
| | | Word fluency | Animals |
| | | Trail Making Test (TMT) | Part A (TRAIL A);<br>Part B (TRAIL B);<br>TRAILBnorm = TRAIL B / TRAIL A |
| | | Boston Naming Test (BNT) | |
| | Apolipoprotein E (APOE) (3) | E2 allele count | |
| | | E3 allele count | |
| | | E4 allele count | |
| | Cognitive Diagnosis (1) | Diagnosis | Healthy control, mild cognitive impairment, or dementia |
| | Health information (9) | Cardio- and cerebrovascular health | |
| | | Diabetes | |
| | | Hypercholesterolemia | |
| | No. sessions before baseline (1) | No. sessions before baseline | |

**Table B (continued)**

| Modality (# features) | Group (# features) | Test/Variables | Information |
|---|---|---|---|
| Structural (35) | Regional grey matter volume (14) | Accumbens | Left and right |
| | | Amygdala | Left and right |
| | | Caudate | Left and right |
| | | Hippocampus | Left and right |
| | | Pallidum | Left and right |
| | | Putamen | Left and right |
| | | Thalamus | Left and right |
| | Global measurements (21) | Total gray matter volume | |
| | | Total subcortical gray matter volume | |
| | | Lateral ventricles | Left and right |
| | | 3rd ventricles | Left and right |
| | | 4th ventricles | Left and right |
| | | Mean cortical thickness | Left and right |
| | | Total cortical volume | Left and right |
| | | Cerebral white matter volume | Left and right |
| | | Cerebellar white matter volume | Left and right |
| | | Cerebellar cortical volume | Left and right |
| | | Corpus callosum | Anterior, mid-anterior, central, mid-posterior and posterior |

*Note*. The following non-brain features were omitted from Vieira et al. (2022) due to excess prevalence of missing values (above 25%): TMT Trail A (RR & LI), TMT Trail B (RR & LI), Word fluency (Vegetables), WMS-R Digit span (DIGIF, DIGIFLEN, DIGIB, DIGIBLEN), Wechsler Adult Intelligence Scale-Revised (WAIS-R) Digit symbol, Family history of dementia (diagnosis of dementia of mother, father and number of siblings diagnosed with dementia), smoking habits (has smoked within the last 30 days, smoked more than 100 cigarettes in their lifetime, total number of years smoking, the average number of cigarette packs smoked per day).

**Table C**

*Comparisons of Absolute Errors for Different Subgroups in Predictions Across Datasets.*

| Dataset | Modality: (1) vs. (2) | CDR-SOB | | | MMSE | | |
|---|---|---|---|---|---|---|---|
| | | *p* | *MAE* (1) | *MAE* (2) | *p* | *MAE* (1) | *MAE* (2) |
| OASIS-3 → ADNI | HC vs. MCI and AD | < .001 | 0.16 | 0.62 | < .001 | 0.31 | 1.1 |
| | MCI vs. HC and AD | < .001 | 0.47 | 0.44 | < .001 | 0.77 | 0.81 |
| | AD vs. HC and MCI | < .001 | 1.2 | 0.33 | < .001 | 2.1 | 0.57 |
| | HC vs. MCI | < .001 | 0.16 | 0.47 | < .001 | 0.31 | 0.77 |
| | HC vs. AD | < .001 | 0.16 | 1.2 | < .001 | 0.31 | 2.1 |
| | MCI vs. AD | < .001 | 0.47 | 1.2 | < .001 | 0.77 | 2.1 |
| | Female vs. Male | < .001 | 0.4 | 0.50 | < .001 | 0.73 | 0.84 |
| | APOE E4: 0 vs. 1 and 2 | < .001 | 0.38 | 0.54 | < .001 | 0.66 | 0.95 |
| | APOE E4: 1 vs. 0 and 2 | .472 | 0.49 | 0.43 | .014 | 0.88 | 0.74 |
| | APOE E4: 2 vs. 0 and 1 | < .001 | 0.71 | 0.42 | < .001 | 1.2 | 0.74 |
| | APOE E4: 0 vs. 1 | .014 | 0.38 | 0.49 | < .001 | 0.66 | 0.88 |
| | APOE E4: 0 vs. 2 | < .001 | 0.38 | 0.71 | < .001 | 0.66 | 1.2 |
| | APOE E4: 1 vs. 2 | < .001 | 0.49 | 0.71 | < .001 | 0.88 | 1.2 |
| | Age: < 65 vs. ≥ 65 years | .838 | 0.41 | 0.46 | .419 | 0.69 | 0.81 |
| | Age: < 70 vs. ≥ 70 years | < .001 | 0.38 | 0.50 | < .001 | 0.67 | 0.87 |
| | Age: < 75 vs. ≥ 75 years | < .001 | 0.41 | 0.53 | < .001 | 0.75 | 0.87 |
| | Age: < 80 vs. ≥ 80 years | < .001 | 0.43 | 0.56 | < .001 | 0.76 | 0.98 |
| ADNI → OASIS-3 | HC vs. MCI and AD | < .001 | 0.19 | 0.69 | < .001 | 0.40 | 0.99 |
| | MCI vs. HC and AD | .014 | 0.66 | 0.30 | > .999 | 0.58 | 0.53 |
| | AD vs. HC and MCI | < .001 | 0.74 | 0.21 | < .001 | 1.1 | 0.42 |
| | HC vs. MCI | < .001 | 0.19 | 0.66 | .939 | 0.40 | 0.58 |
| | HC vs. AD | < .001 | 0.19 | 0.74 | < .001 | 0.40 | 1.1 |
| | MCI vs. AD | .841 | 0.66 | 0.74 | .731 | 0.58 | 1.1 |
| | Female vs. Male | < .001 | 0.27 | 0.34 | < .001 | 0.48 | 0.60 |
| | APOE E4: 0 vs. 1 and 2 | .009 | 0.26 | 0.37 | .398 | 0.47 | 0.64 |
| | APOE E4: 1 vs. 0 and 2 | .087 | 0.37 | 0.27 | .758 | 0.63 | 0.49 |
| | APOE E4: 2 vs. 0 and 1 | .051 | 0.40 | 0.30 | .939 | 0.70 | 0.52 |
| | APOE E4: 0 vs. 1 | .048 | 0.26 | 0.37 | .670 | 0.47 | 0.63 |
| | APOE E4: 0 vs. 2 | .015 | 0.26 | 0.40 | .758 | 0.47 | 0.70 |
| | APOE E4: 1 vs. 2 | .336 | 0.37 | 0.40 | > .999 | 0.63 | 0.70 |
| | Age: < 65 vs. ≥ 65 years | < .001 | 0.17 | 0.33 | .015 | 0.36 | 0.57 |
| | Age: < 70 vs. ≥ 70 years | < .001 | 0.21 | 0.38 | .018 | 0.43 | 0.62 |
| | Age: < 75 vs. ≥ 75 years | < .001 | 0.23 | 0.46 | .006 | 0.46 | 0.70 |
| | Age: < 80 vs. ≥ 80 years | < .001 | 0.27 | 0.51 | .024 | 0.49 | 0.75 |

*Note*. The *p* values of comparisons of absolute errors for different subgroups in predictions across datasets are displayed. (1) refers to the left side of the comparison, (2) refers to the right side of the comparison. Two-sided Mann-Whitney U tests ($\alpha = .05$) for comparison of absolute errors were performed. *p* values were corrected for multiple comparisons using the Holm-Bonferroni method. OASIS-3 → ADNI = model trained on OASIS-3 and tested on ADNI; ADNI → OASIS-3 = model trained on ADNI and tested on OASIS-3. HC = healthy control; MCI = mild cognitive impairment, AD = Alzheimer's disease or another form of dementia; APOE = apolipoprotein E; *MAE* = mean absolute error.

**Table D**

*Permutation Importance of All Features of OASIS-3 and ADNI.*

| OASIS-3 | | ADNI | |
|---|---|---|---|
| Feature | PI | Feature | PI |
| mmse_mmse | 0.072 (0.069, 0.075) | TMTB_TRAILB | 0.040 (0.038, 0.042) |
| WMSlm_MEMUNITS | 0.071 (0.067, 0.075) | mmse_mmse | 0.031 (0.030, 0.032) |
| faq_faqSum | 0.053 (0.049, 0.057) | WMSlm_MEMUNITS | 0.030 (0.028, 0.031) |
| Vol. of Left-Hippocampus | 0.027 (0.026, 0.029) | faq_BILLS | 0.013 (0.012, 0.014) |
| WMSlm_LOGIMEM | 0.018 (0.016, 0.019) | WMSlm_LOGIMEM | 0.012 (0.011, 0.013) |
| TMTB_TRAILB | 0.013 (0.012, 0.014) | TMTA_TRAILA | 0.0090 (0.0075, 0.011) |
| cdr_sob | 0.0068 (0.0050, 0.0086) | faq_faqSum | 0.0075 (0.0061, 0.0088) |
| Vol. of Left-Amygdala | 0.0057 (0.0043, 0.0070) | BOSTON_BOSTON | 0.0062 (0.0053, 0.0071) |
| WF_ANIMALS | 0.0050 (0.0041, 0.0058) | WF_ANIMALS | 0.0061 (0.0043, 0.0079) |
| Vol. of Right-Accumbens-area | 0.0045 (0.0039, 0.0051) | lh_MeanThickness_thickness | 0.0061 (0.0055, 0.0066) |
| Vol. of CC_Central | 0.0043 (0.0035, 0.0050) | npiq_npiqSevSum | 0.0048 (0.0042, 0.0054) |
| Vol. of Right-Amygdala | 0.0040 (0.0032, 0.0049) | demo_age | 0.0039 (0.0035, 0.0044) |
| TMTB_TRAILBnorm | 0.0037 (0.0032, 0.0042) | Vol. of Right-Hippocampus | 0.0037 (0.0034, 0.0041) |
| Vol. of Right-Hippocampus | 0.0037 (0.0029, 0.0044) | Vol. of 4th-Ventricle | 0.0036 (0.0032, 0.0040) |
| Vol. of Right-Thalamus-Proper | 0.0032 (0.0026, 0.0039) | faq_TAXES | 0.0030 (0.0024, 0.0037) |
| faq_REMDATES | 0.0031 (0.0024, 0.0037) | Vol. of Left-Amygdala | 0.0027 (0.0023, 0.0032) |
| Vol. of Left-Pallidum | 0.0025 (0.0021, 0.0030) | TMTB_TRAILBnorm | 0.0024 (0.0018, 0.0030) |
| lhCerebralWhiteMatterVol | 0.0021 (0.0014, 0.0027) | Vol. of 3rd-Ventricle | 0.0021 (0.0016, 0.0026) |
| Vol. of Left-Cerebellum-White-Matter | 0.0019 (0.0015, 0.0023) | lhCortexVol | 0.0015 (0.00095, 0.0021) |
| npiq_npiqPresSum | 0.0018 (0.0015, 0.0022) | Vol. of Left-Accumbens-area | 0.0014 (0.00086, 0.0019) |
| Vol. of Left-Accumbens-area | 0.0018 (0.0012, 0.0024) | faq_EVENTS | 0.0013 (0.00091, 0.0017) |
| WMSlm_MEMTIME | 0.0014 (0.0011, 0.0018) | rhCortexVol | 0.0013 (0.00057, 0.0020) |
| cdr_homehobb | 0.0014 (0.00077, 0.0020) | cdr_sob | 0.0012 (0.00066, 0.0018) |
| cvasc_HYPERTEN | 0.0012 (0.00093, 0.0015) | cdr_memory | 0.0012 (0.00072, 0.0017) |
| Vol. of Right-Cerebellum-White-Matter | 0.0012 (0.00072, 0.0016) | Vol. of Left-Hippocampus | 0.0012 (0.00094, 0.0014) |
| cdr_commun | 0.00096 (0.00061, 0.0013) | Vol. of Right-Amygdala | 0.00097 (0.00066, 0.0013) |
| npiq_npiqSevSum | 0.00073 (0.00042, 0.0010) | Vol. of Right-Accumbens-area | 0.00092 (0.00056, 0.0013) |
| Vol. of 4th-Ventricle | 0.00071 (0.00036, 0.0011) | Vol. of Left-Pallidum | 0.00067 (0.00041, 0.00094) |
| TMTA_TRAILA | 0.00065 (0.00018, 0.0011) | faq_REMDATES | 0.00054 (0.00013, 0.00095) |

### Table D (continued)

| OASIS-3 | | ADNI | |
|---|---|---|---|
| Feature | PI | Feature | PI |
| apoe_e4count | 0.00058 (0.00040, 0.00076) | WMSlm_MEMTIME | 0.00047 (0.00025, 0.00069) |
| Vol. of Right-Putamen | 0.00053 (0.00019, 0.00087) | cvasc_HYPERTEN | 0.00043 (0.00030, 0.00056) |
| cdr_cdrGlobal | 0.00051 (0.00013, 0.00089) | apoe_e4count | 0.00040 (0.00027, 0.00053) |
| Vol. of Right-Lateral-Ventricle | 0.00048 (0.00013, 0.00082) | Vol. of CC_Central | 0.00037 (0.00015, 0.00060) |
| hypercho_HYPERCHO | 0.00041 (0.00020, 0.00063) | rhCerebralWhiteMatterVol | 0.00031 (-0.000039, 0.00066) |
| Vol. of Left-Putamen | 0.00038 (-0.000067, 0.00083) | Vol. of Left-Thalamus-Proper | 0.00031 (0.000036, 0.00058) |
| Vol. of SubCortGrayVol | 0.00033 (0.000049, 0.00062) | npiq_npiqPresSum | 0.00027 (-0.000074, 0.00062) |
| Vol. of CC_Mid_Anterior | 0.00031 (0.00000011, 0.00062) | Vol. of SubCortGrayVol | 0.00021 (-0.0000016, 0.00041) |
| diag | 0.00019 (-0.00064, 0.0010) | rh_MeanThickness_thickness | 0.00019 (-0.000087, 0.00046) |
| demo_sex | 0.00015 (0.0000030, 0.00029) | Vol. of CC_Anterior | 0.00016 (-0.000067, 0.00039) |
| Vol. of CC_Anterior | 0.000090 (-0.00025, 0.00043) | Vol. of CC_Mid_Posterior | 0.00016 (-0.000064, 0.00039) |
| diabetes_DIABETES | 0.000065 (-0.000044, 0.00017) | demo_education | 0.00016 (0.0000022, 0.00032) |
| session_n | 0.000032 (-0.00013, 0.00020) | faq_GAMES | 0.00015 (-0.00021, 0.00051) |
| Vol. of Left-Lateral-Ventricle | 0.000031 (-0.00044, 0.00050) | lhCerebralWhiteMatterVol | 0.00014 (-0.00012, 0.00040) |
| cdr_judgment | 0.000010 (-0.00024, 0.00026) | cvasc_CVAFIB | 0.00013 (0.00010, 0.00015) |
| apoe_e3count | 0.0000041 (-0.00015, 0.00016) | demo_sex | 0.00010 (-0.00000093, 0.00021) |
| cdr_perscare | -0.000028 (-0.00011, 0.000053) | apoe_e3count | 0.000072 (-0.000033, 0.00018) |
| cvasc_CVHATT | -0.000051 (-0.00012, 0.000018) | Vol. of Left-Caudate | 0.000059 (-0.00014, 0.00026) |
| faq_BILLS | -0.000077 (-0.00029, 0.00013) | cdr_orient | 0.000056 (-0.00012, 0.00023) |
| cvasc_CVOTHR | -0.000077 (-0.00016, 0.0000086) | session_n | 0.000040 (0.0000054, 0.000075) |
| faq_STOVE | -0.000080 (-0.00017, 0.000014) | Vol. of Right-Thalamus-Proper | 0.000038 (-0.00021, 0.00029) |
| Vol. of Left-Caudate | -0.00010 (-0.00042, 0.00021) | apoe_e2count | 0.000024 (-0.000020, 0.000068) |
| apoe_e2count | -0.00010 (-0.00017, -0.000038) | Vol. of Right-Cerebellum-Cortex | 0.0000031 (-0.00018, 0.00019) |
| demo_education | -0.00013 (-0.00065, 0.00040) | cvasc_CVHATT | 0.00000025 (-0.0000044, 0.0000049) |
| Vol. of Right-Pallidum | -0.00016 (-0.00039, 0.000070) | cvasc_CBSTROKE | -0.000016 (-0.000032, 0.00000022) |
| cvasc_CVAFIB | -0.00017 (-0.00024, -0.000099) | cdr_judgment | -0.000019 (-0.00020, 0.00016) |
| cvasc_CVANGIO | -0.00018 (-0.00030, -0.000051) | cvasc_CBTIA | -0.000030 (-0.000053, -0.0000081) |
| Vol. of Left-Thalamus-Proper | -0.00018 (-0.00041, 0.000050) | cdr_homehobb | -0.000032 (-0.00036, 0.00030) |
| Vol. of Left-Cerebellum-Cortex | -0.00021 (-0.00051, 0.000076) | Vol. of Left-Cerebellum-White-Matter | -0.000033 (-0.00033, 0.00026) |

**Table D (continued)**

| OASIS-3 | | ADNI | |
|---|---|---|---|
| Feature | PI | Feature | PI |
| Vol. of 3rd-Ventricle | -0.00022 (-0.00054, 0.000095) | cdr_cdrGlobal | -0.000034 (-0.00020, 0.00013) |
| faq_GAMES | -0.00023 (-0.00047, 0.0000058) | cdr_perscare | -0.000045 (-0.00014, 0.000053) |
| Vol. of CC_Mid_Posterior | -0.00026 (-0.00092, 0.00039) | hypercho_HYPERCHO | -0.000056 (-0.00013, 0.000021) |
| faq_MEALPREP | -0.00028 (-0.00046, -0.00011) | diabetes_DIABETES | -0.000064 (-0.00012, -0.0000074) |
| Vol. of Right-Cerebellum-Cortex | -0.00030 (-0.00054, -0.000055) | Vol. of Left-Cerebellum-Cortex | -0.00011 (-0.00029, 0.000073) |
| faq_PAYATTN | -0.00033 (-0.00052, -0.00014) | faq_PAYATTN | -0.00012 (-0.00048, 0.00024) |
| faq_SHOPPING | -0.00034 (-0.00053, -0.00015) | diag | -0.00012 (-0.00040, 0.00016) |
| faq_TRAVEL | -0.00042 (-0.00072, -0.00011) | cdr_commun | -0.00017 (-0.00037, 0.000038) |
| cvasc_CBSTROKE | -0.00052 (-0.00063, -0.00041) | cvasc_CVOTHR | -0.00019 (-0.00025, -0.00013) |
| faq_TAXES | -0.00053 (-0.00089, -0.00017) | cvasc_CVANGIO | -0.00019 (-0.00029, -0.000092) |
| gds_gdsSum | -0.00054 (-0.00078, -0.00030) | faq_MEALPREP | -0.00020 (-0.00058, 0.00018) |
| rhCortexVol | -0.00058 (-0.0011, -0.0000075) | Vol. of Right-Pallidum | -0.00022 (-0.00042, -0.000015) |
| cvasc_CBTIA | -0.00061 (-0.00087, -0.00035) | gds_gdsSum | -0.00032 (-0.00047, -0.00017) |
| cdr_orient | -0.00065 (-0.00093, -0.00038) | Vol. of Left-Putamen | -0.00034 (-0.00053, -0.00014) |
| rhCerebralWhiteMatterVol | -0.00068 (-0.0011, -0.00027) | Vol. of Right-Cerebellum-White-Matter | -0.00036 (-0.00060, -0.00012) |
| demo_age | -0.00070 (-0.0012, -0.00020) | Vol. of Right-Putamen | -0.00043 (-0.00065, -0.00020) |
| Vol. of Right-Caudate | -0.00079 (-0.0011, -0.00048) | Vol. of CC_Mid_Anterior | -0.00046 (-0.00069, -0.00022) |
| faq_EVENTS | -0.00084 (-0.0011, -0.00060) | Vol. of Right-Caudate | -0.00053 (-0.00071, -0.00035) |
| lh_MeanThickness_thickness | -0.00087 (-0.0015, -0.00024) | Vol. of TotalGrayVol | -0.00061 (-0.00093, -0.00030) |
| Vol. of TotalGrayVol | -0.00095 (-0.0016, -0.00030) | faq_STOVE | -0.00066 (-0.00087, -0.00045) |
| Vol. of CC_Posterior | -0.0010 (-0.0012, -0.00082) | Vol. of Left-Lateral-Ventricle | -0.00078 (-0.0010, -0.00056) |
| cdr_memory | -0.0011 (-0.0014, -0.00070) | faq_SHOPPING | -0.00089 (-0.0012, -0.00056) |
| rh_MeanThickness_thickness | -0.0017 (-0.0021, -0.0014) | Vol. of CC_Posterior | -0.00098 (-0.0012, -0.00078) |
| BOSTON_BOSTON | -0.0022 (-0.0028, -0.0016) | faq_TRAVEL | -0.0011 (-0.0014, -0.00074) |
| lhCortexVol | -0.0033 (-0.0042, -0.0024) | Vol. of Right-Lateral-Ventricle | -0.0012 (-0.0015, -0.00086) |

*Note*. Permutation importance of all input features of the models including non-brain data and structural MRI data for prediction within dataset, ordered by importance. PI = median permutation importance across 1000 splits.

**Table E**

*Overview of Model Performance Including Post hoc Analysis With Matched Samples.*

| Dataset | Modality | CDR-SOB | | | MMSE | | |
|---|---|---|---|---|---|---|---|
| | | $R^2$ | *MSE* | *MAE* | $R^2$ | *MSE* | *MAE* |
| OASIS-3 → ADNI | non-brain | .30 | .65 | .50 | .25 | 1.99 | .81 |
| | structural MRI | .17 | .77 | .49 | .10 | 2.37 | .89 |
| | combined | .35 | .60 | .45 | .26 | 1.95 | .79 |
| | combined *matched* | .22 | .36 | .38 | .18 | .84 | .56 |
| | top 15 | .34 | .61 | .42 | .26 | 1.96 | .80 |
| | top 15 *matched* | .27 | .34 | .33 | .16 | .87 | .57 |
| ADNI → OASIS-3 | non-brain | .28 | .27 | .29 | .19 | .71 | .48 |
| | structural MRI | −.02 | .38 | .41 | −.15 | 1.00 | .68 |
| | combined | .31 | .26 | .30 | .18 | .72 | .53 |
| | combined *matched* | .29 | .42 | .42 | .29 | 1.1 | .68 |
| | top 15 | .31 | .26 | .27 | .18 | .71 | .47 |
| | top 15 *matched* | .29 | .42 | .39 | .25 | 1.2 | .65 |

*Note*. This Table is an extended and adapted version of Table 2, including the post hoc analysis with matched samples, excluding the model comparisons. *matched* indicates that the distribution of the predictive targets (CDR-SOB and MMSE slope) of the test set was matched to that of the training set in predictions across datasets. OASIS-3 → ADNI = model trained on OASIS-3 and tested on ADNI; ADNI → OASIS-3 = model trained on ADNI and tested on OASIS-3; CDR-SOB = Clinical Dementia Rating Scale Sum of Boxes; MMSE = Mini-Mental State Examination; $R^2$ = median coefficient of determination across 1000 splits; *MSE* = mean squared error; *MAE* = mean absolute error.

### B. Supplementary Figures

#### Figure A

*Inclusion of Subjects of the ADNI Dataset.*

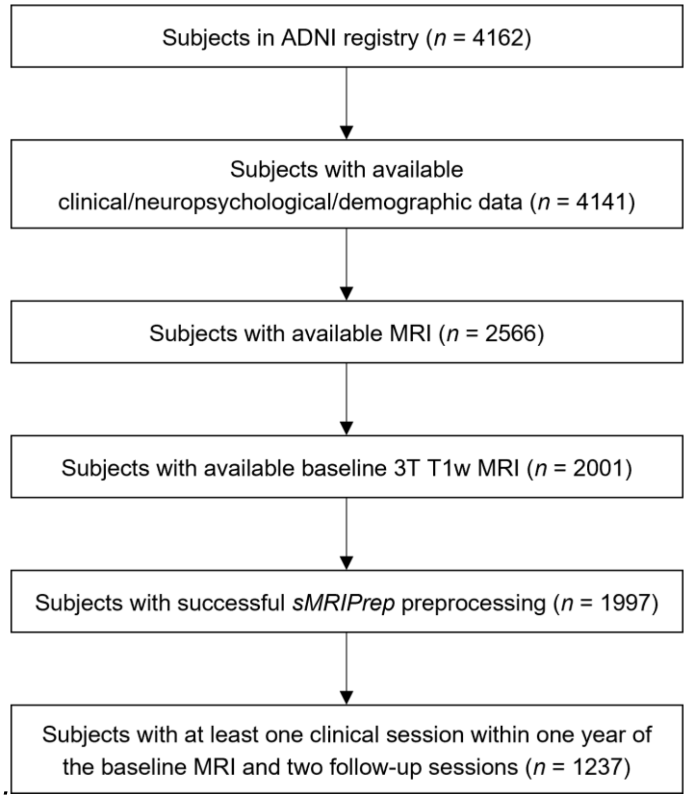

*Note*. The final ADNI dataset consisted of 1237 participants. T1w MRI = T1-weighted magnetic resonance imaging.

**Figure B**

*Distribution of CDR-SOB and MMSE Change in OASIS-3 and ADNI.*

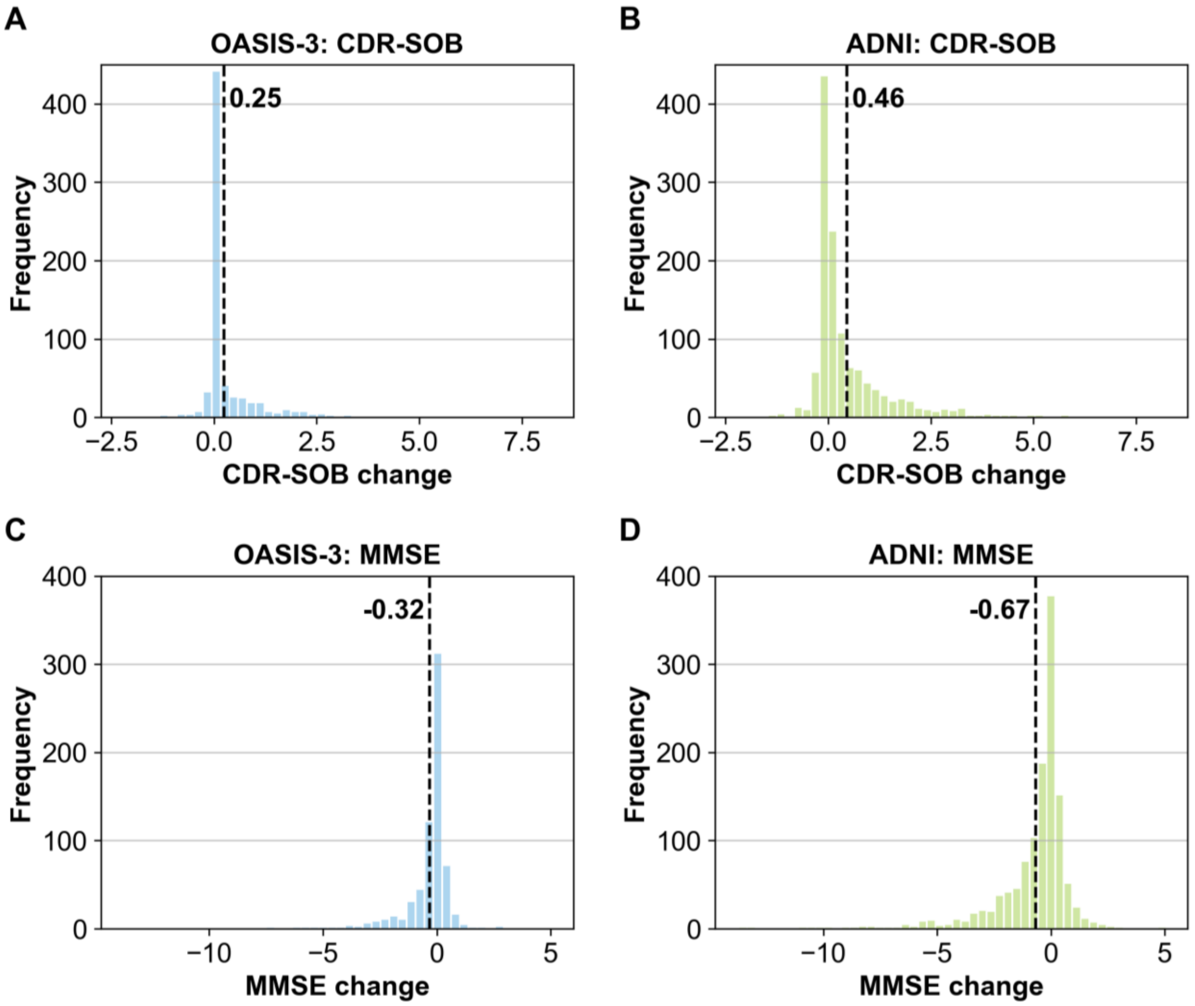

*Note*. Panels A and C: Distribution of observed cognitive decline measured via Clinical Dementia Rating Scale Sum of Boxes (CDR-SOB change) and Mini-Mental State Examination (MMSE change) in the OASIS-3 dataset. Panels B and D: Distribution of observed CDR-SOB change and MMSE change in the ADNI dataset. CDR-SOB change: Positive values indicate cognitive decline. MMSE change: Negative values indicate cognitive decline. The number and the dashed vertical line represent the mean cognitive change.

**Figure C**

*Distribution of $R^2$ Across 1000 Splits in OASIS-3 Models.*

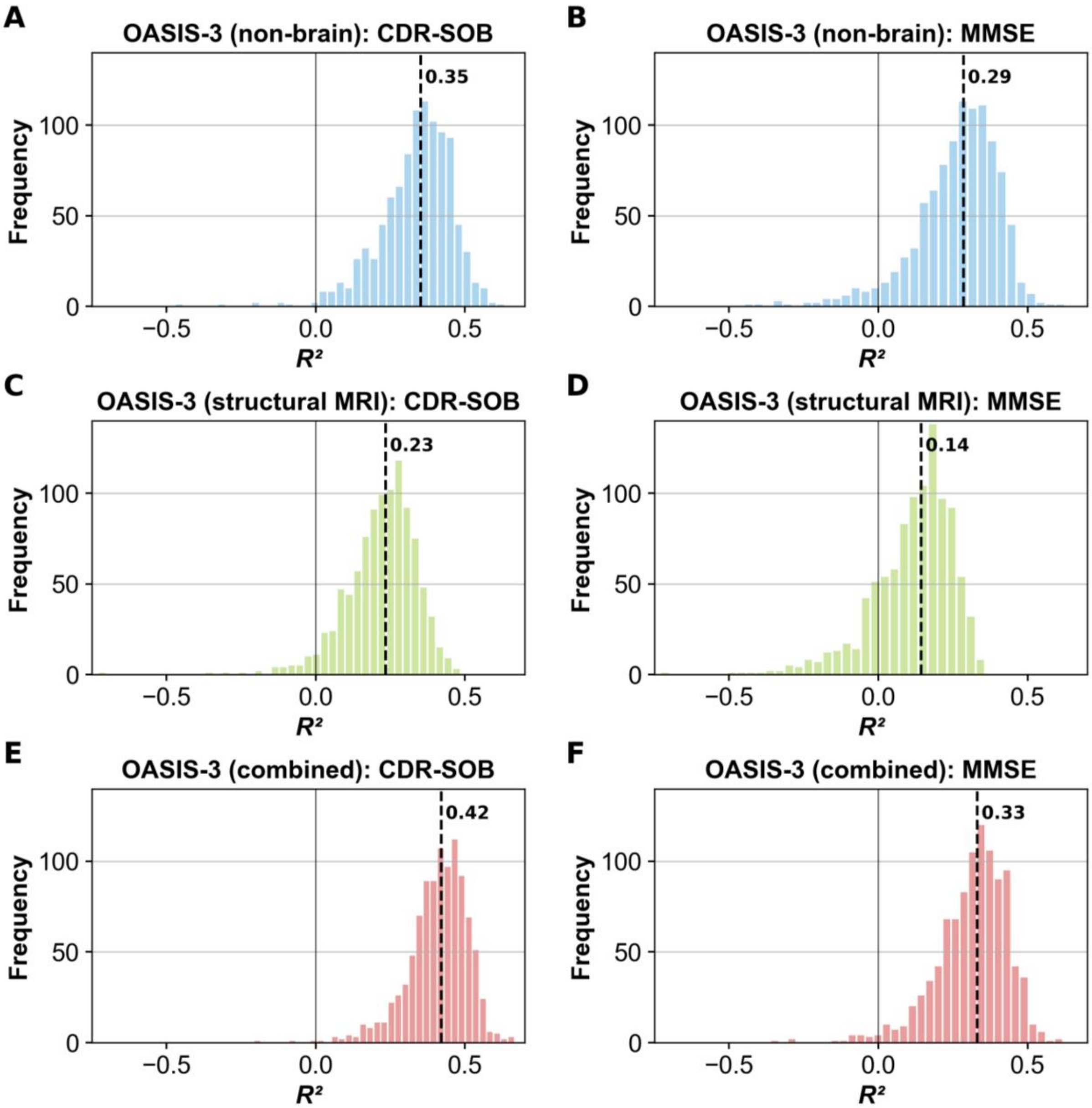

*Note*. Distribution of $R^2$ (coefficient of determination) across 1000 splits for predicting cognitive decline measured via Clinical Dementia Rating Scale Sum of Boxes (CDR-SOB change) (in panels A, B and C) and Mini-Mental State Examination (MMSE change) (in panels D, E and F) when using different input modalities (non-brain; structural MRI; combined) in OASIS-3 data. The number and the dashed vertical line represent the median $R^2$ across 1000 splits.

**Figure D**

*Distribution of $R^2$ Across 1000 Splits in ADNI Models.*

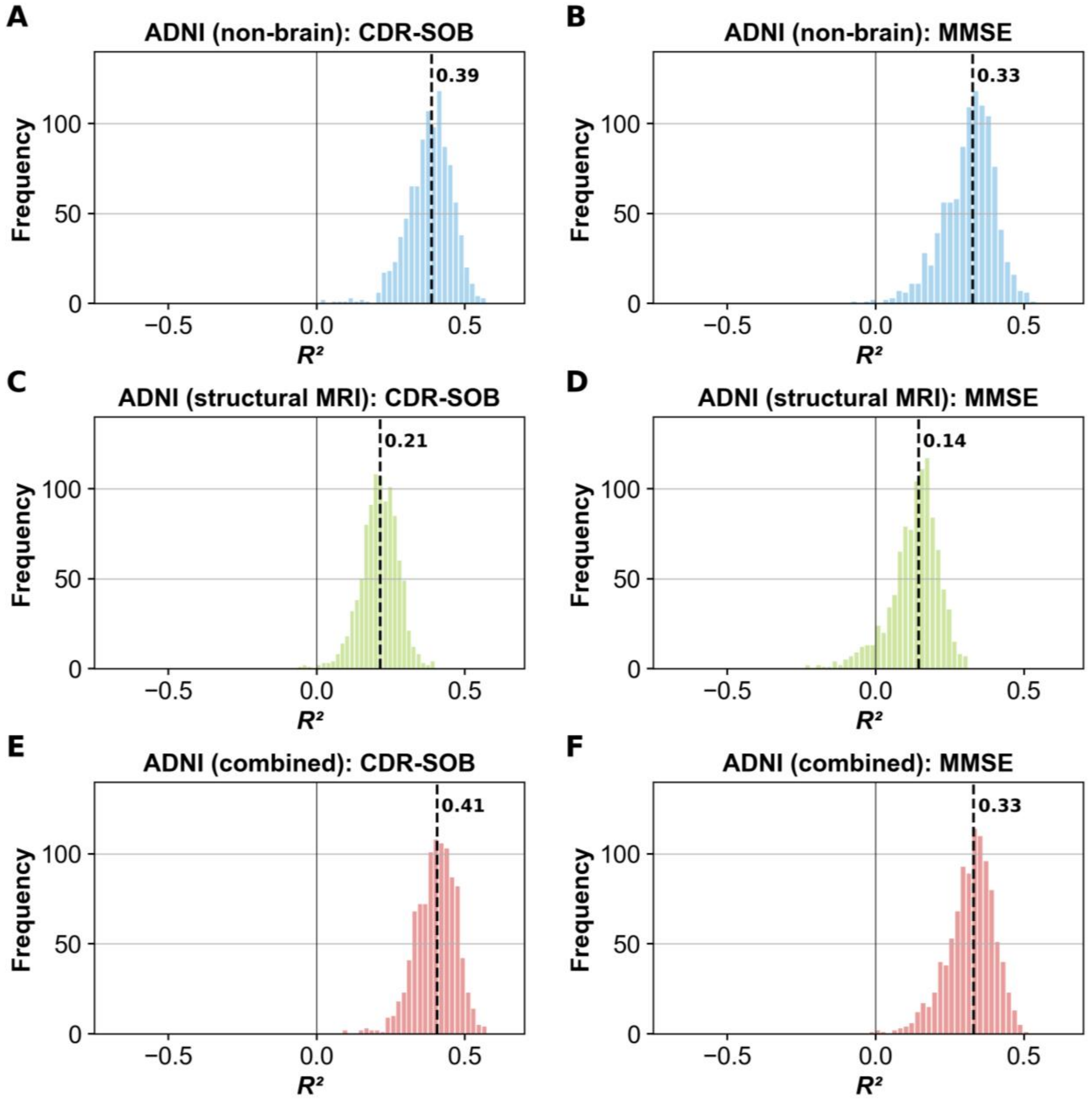

*Note*. Distribution of $R^2$ (coefficient of determination) across 1000 splits for predicting cognitive decline measured via Clinical Dementia Rating Scale Sum of Boxes (CDR-SOB change) (in panels A, B and C) and Mini-Mental State Examination (MMSE change) (in panels D, E and F) when using different input modalities (non-brain; structural MRI; combined) in ADNI data. The number and the dashed vertical line represent the median $R^2$ across 1000 splits.

**Figure E**

*True Versus Predicted Values of CDR-SOB and MMSE Change in Predictions Across Datasets.*

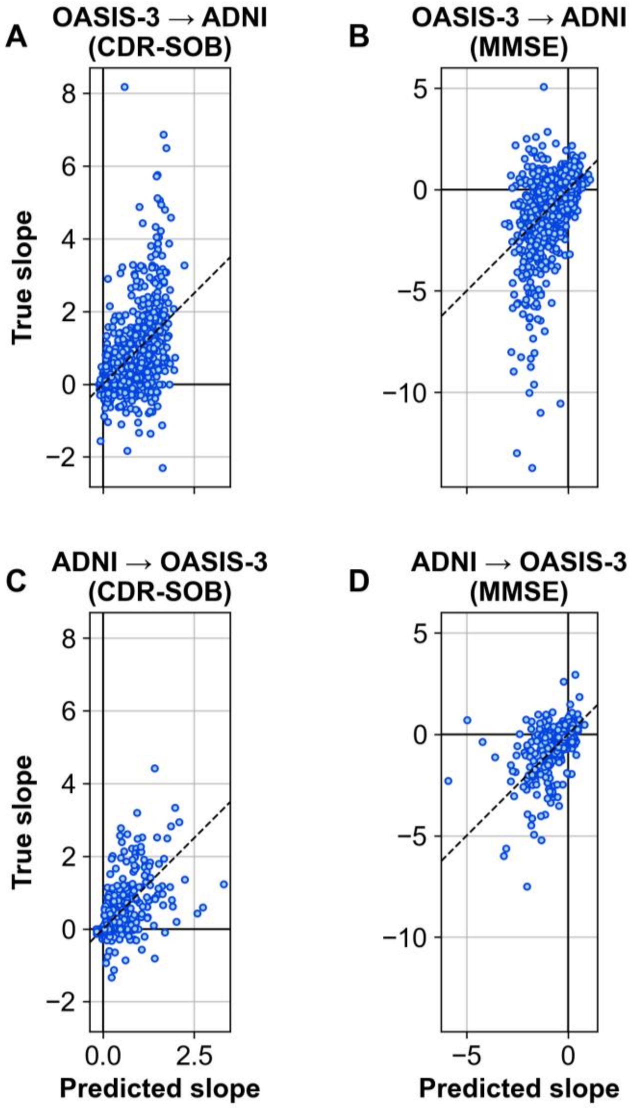

*Note*. Combined models included non-brain data and structural MRI data to predict continuous cognitive decline across datasets. The scatter plot shows true versus predicted values of CDR-SOB change (A, C) or MMSE change (B, D) for models trained on OASIS-3 and tested on ADNI (A, B) and models trained on ADNI and tested on OASIS-3 (C, D). CDR-SOB change: Positive values indicate cognitive decline. MMSE change: Negative values indicate cognitive decline. OASIS-3 → ADNI = model trained on OASIS-3 and tested on ADNI; ADNI → OASIS-3 = model trained on ADNI and tested on OASIS-3.

**Figure F**

*True Versus Predicted Values of CDR-SOB and MMSE Change for Subgroups Differentiated by Clinical Diagnoses in Predictions Across Datasets.*

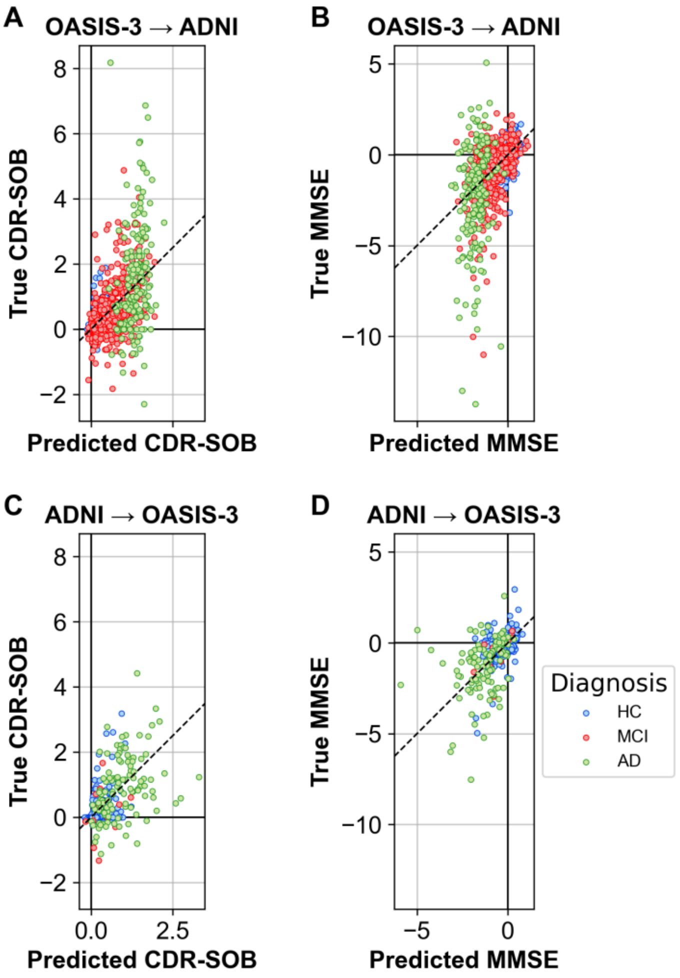

*Note*. Combined models included non-brain data and structural MRI data to predict continuous cognitive decline across datasets. The scatter plot shows true versus predicted values of CDR-SOB and MMSE change. CDR-SOB change: Positive values indicate cognitive decline. MMSE change: Negative values indicate cognitive decline. Each data point represents an individual participant of the respective test set. Data points are differentiated regarding the clinical diagnoses of the participants in the test set. HC = healthy control; MCI = mild cognitive impairment, AD = Alzheimer's disease or another form of dementia; OASIS-3 → ADNI = model trained on OASIS-3 and tested on ADNI; ADNI → OASIS-3 = model trained on ADNI and tested on OASIS-3. 30 OASIS-3 participants with missing diagnoses are omitted in C and D.

### Figure G

*Feature Importance for Predictions Across Datasets*

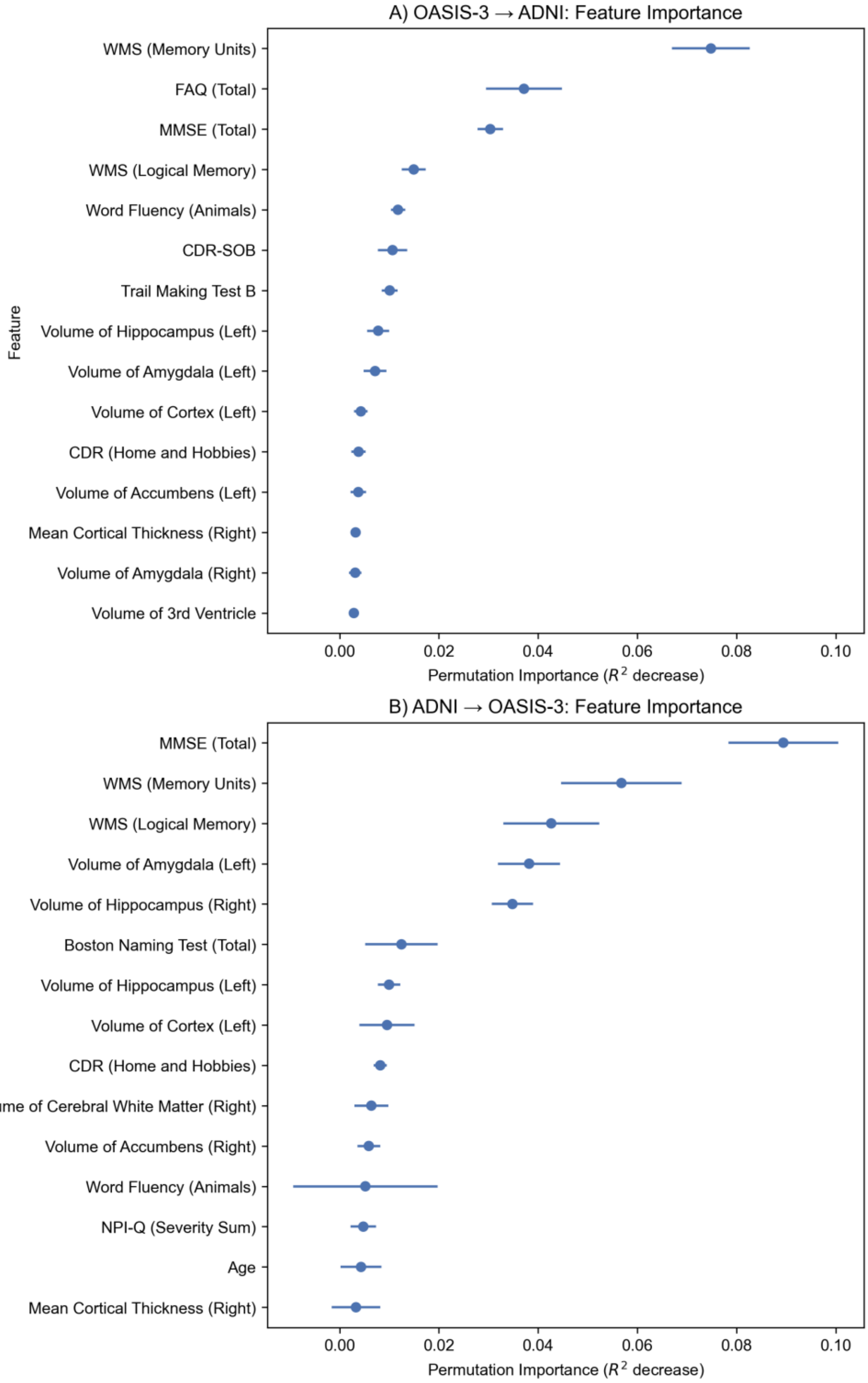

*Note*. Permutation importance of the top 15 features of the combined models for prediction across dataset with 100 repetitions, ordered by average importance.
